# Why Have Drug Names Become So Weird? A Retrospective Analysis of the Orthographic Distinctiveness of Proprietary and Generic Names Across Four Decades of FDA Approvals

**DOI:** 10.64898/2026.10.15.26365645

**Authors:** Bruce L. Lambert

**Affiliations:** Department of Communication Studies, Northwestern University, Evanston, IL, USA

**Keywords:** drug names, medication errors, drug name confusion, n-gram analysis, FDA, proprietary names

## Abstract

**Background:** Regulatory pressure to avoid confusingly similar drug names appears to have pushed manufacturers to coin more distinctive proprietary names, but no one has quantitatively analyzed this trend.

**Objective:** To determine whether proprietary drug names became more orthographically distinctive (unusual in spelling) over time.

**Methods:** We examined 1,321 US Food and Drug Administration (FDA) new molecular entity (NME) names (1985–2024), 2,769 FDA Orange Book trade names (1982–2026), 177,337 US pharmaceutical trademarks (1980–2026), and 1,273 generic names (1982–2025). Compared to prior names (including those already marketed) and ordinary English, we scored each name’s character trigram (three-letter sequence) surprisal (statistical unexpectedness) and its edit-distance similarity to the closest existing names. Linear regression tested time trends, controlling for name length.

**Results:** Mean NME trigram surprisal rose in every decade between the 1980s and 2020s, from 7.21 to 8.50 nats (natural-logarithm units, higher meaning less predictable), an 18% increase (b = 0.038, p < .001, R² = 0.34). Half of 2020s names (7% in the 1980s) contained a letter sequence found in no common English word (e.g., *Jesduvroq*, *Omvoh*). Nearest-neighbor similarity fell after the early 1990s. Trademark data showed the same trend (b = 0.016, p < .001). Generic names showed a weaker trend (b = 0.011, R² = 0.063).

**Conclusions:** Over four decades, drug names became more orthographically distinctive, consistent with namespace crowding, deliberate efforts to avoid confusing names, and branding strategy. Because unfamiliar letter sequences can introduce other error pathways, increased distinctiveness does not necessarily imply improved safety.

## Introduction

In the 1980s and 1990s, the FDA approved drug names like *Cardene*, *Claritin*, *Serevent*, and *Vantin*. In the 2020s it approved *Zynyz*, *Xolremdi*, *Bimzelx*, *Zurzuvae*, *Jesduvroq*, and *Omvoh*. The older names resembled English words. In contrast, newer names have letter sequences that never occur in common English words (e.g., *omv*, *mvo*, and *voh* in *Omvoh*) Distinctive spelling might reduce drug name confusion, but it might also make names harder to read, pronounce, and remember. This study begins with the subjective impression that modern drug names have become stranger over time and tests it with quantitative analysis.

Choosing a proprietary (brand) drug name is an important commercial decision. If a name is too similar to an existing product, it can cause dispensing errors, prescribing errors, or patient harm [1–3]. Name confusion is often said to cause up to 25% of medication errors [1, 2], but those figures come from error *reports* not observed error *rates*. In a national direct-observation study of 4,481 prescriptions in 50 US pharmacies, 6 (0.13%) were filled with the wrong drug [4]. A similar rate (0.14%) was found when indication alerts intercepted drug name confusion errors during computerized order entry (176 of 127,458 alerted orders) [5]. Both rates are near 1 in 1,000. A US Pharmacopeia review of two national reporting programs (2003–2006) counted 26,604 reported look-alike or sound-alike errors involving more than 3,170 name pairs, but did not give a rate [6].

The World Health Organization (WHO) highlights look-alike, sound-alike (LASA) medication errors in technical guidance for its Third Global Patient Safety Challenge [7], and the Institute for Safe Medication Practices (ISMP) maintains a list of over 500 confused drug name pairs [8]. Before approval, the US Food and Drug Administration (FDA) reviews proposed proprietary names for safety concerns (including confusing similarity) [9].

Experimental studies confirm the link between name similarity and confusion. When pharmacists see degraded drug names, they are more likely to misidentify names that have many close orthographic (spelling) neighbors [10]. Orthographic and phonological (sound) similarity also increase false recognition of drug names in memory tasks [11], and similarity affects short-term memory for drug names in systematic ways (though the effects vary by task and stimulus characteristics) [12]. Thus, orthographic distinctiveness is not just a theoretical concern but also a potential risk factor for confusion errors.

As the lexicon of drug names grows, the available namespace for new names shrinks, because each new approval must be sufficiently distinct from all other existing names. Industry observers note that tens of thousands of existing trademarks make distinctive names hard to coin [13]. In letter-frequency analysis, proprietary names diverge from English, with *X*, *Z*, and *V* markedly overrepresented and *V*, *Y*, and *Z* increasingly common [14]. The pattern parallels broader trademark congestion, where the availability of effective word marks is finite and shrinking [15].

Many have commented on the subjective sense that drug names are increasingly unusual in their spelling, but to our knowledge no one has quantified this empirically in a systematic way across a large set of FDA-approved names over a prolonged period. Several factors influence how drugs are named, including regulatory scrutiny, commercial trends, and the increasing crowding of the trademark namespace [15]. This study cannot isolate any of those causes. What it does do is examine whether the spelling of drug names has become more distinctive over time.

We hypothesized that approved brand names have become more orthographically distinctive from 1985 to 2024. We measured distinctiveness as the rarity of each name’s character trigrams (three-letter sequences) relative to previously approved names. The metric has psycholinguistic grounding, because the frequency of sub-word segments affects spoken word recognition [16]. Rare trigrams do not guarantee low confusability (such a name can still resemble some individual existing name), so we also measured similarity to the closest existing names.

Generic names (International Nonproprietary Names and United States Adopted Names, INN/USAN) face the same pressure, but the WHO INN stem system limits variability by requiring related drugs to share common suffixes (e.g., *-tinib* for kinase inhibitors, *-sartan* for angiotensin receptor blockers). In a secondary analysis, we tested whether generic names show a similar pattern.

Character trigrams are long enough to capture the prefix, suffix, and consonant-cluster patterns that drive visual similarity, and still short enough to avoid sparsity in small data sets. We did not measure phonological (sound-alike) similarity [3, 17].

## Methods

We conducted a retrospective observational study of naming trends, drawing on three publicly available datasets of proprietary drug names and pharmaceutical trademarks (a fourth dataset, of generic names, served as a comparison). Our primary outcome was drug baseline character trigram *surprisal* (how unusual or surprising a name’s letter sequences are relative to all previously approved names). Secondary outcomes were nearest-neighbor edit-distance similarity and the proportion of trigrams absent from ordinary English (what we termed the *novel fraction*). Because all data are publicly available from federal agencies and contain no patient-level information, ethics approval was not required.

### Data Sources

#### Primary dataset (NME compilation)

The main dataset was the FDA Center for Drug Evaluation and Research Compilation of New Molecular Entity (NME) Drug and New Biologic Approvals, a complete record of new drug approvals from 1985 through 2024. We downloaded it programmatically in February 2026 (see Data Availability). We normalized each name by converting it to lower case, removing trademark symbols, punctuation, and digits, and stripping dosage-form suffixes (such as XR and ER). We excluded placeholder entries.

There were 1,321 unique proprietary names in our database. The unit of analysis was the individual unique drug name. We computed trigrams within words, meaning they did not span different words in a multi-word name. The surprisal score for an individual name was the mean of the surprisal scores of each of its trigrams (a trigram occurring twice in a name counted twice). The length of a name was the total number of letters. Edit distances compared whole names, with a single space between the words of a multi-word name (e.g., *breo ellipta*).

#### Replication dataset (FDA Orange Book)

We repeated the analysis on the FDA Orange Book Products file (downloaded February 2026), which covers the broader set of products approved under new drug applications (NDAs). We excluded abbreviated new drug application entries (ANDA, generic). Products approved before 1982 have no approval date in the Orange Book (the date field reads “Approved Prior to Jan 1, 1982”). We therefore scored only trade names first approved in 1982 or later and used the earlier names as the starting baseline (see below). Some Orange Book entries are not coined trade names. We excluded product descriptions (e.g., *potassium chloride in dextrose and sodium chloride in plastic container*) and entries whose trade name is simply the established (generic) name (e.g., *diltiazem hydrochloride*) from scoring and from every baseline (Supplementary Methods). That left 2,769 unique trade names first used in 1982 or later (147 reused pre-1982 names were not counted as new).

#### Replication dataset (USPTO pharmaceutical trademarks)

To test whether the trend extended to proposed names, we extracted International Class 005 (Pharmaceuticals) trademarks from United States Patent and Trademark Office (USPTO) bulk data, keeping marks whose goods and services description contained pharmaceutical keywords and excluding veterinary-only marks (Supplementary Methods). We used the filing date in place of the approval date, kept filings from January 1, 1980, through February 9, 2026 (the last filing date in the file) and names of 3 to 30 characters, and included all statuses (live, dead, pending). The 177,337 unique marks (134 times the size of the NME dataset) therefore include names that were never marketed.

#### Comparison dataset (generic INN/USAN names)

We also extracted the *ingredient* names of each Orange Book NDA product. A combination product contributed each of its constituent ingredient names, and its complete list of ingredients was not itself scored as a separate, multiword name. Biologics, including monoclonal antibodies, are licensed under biologics license applications and are not in the Orange Book NDA file. We removed salt and ester suffixes (dexamethasone sodium phosphate became dexamethasone) and excluded radiopharmaceuticals and simple salts (Supplementary Methods). Ingredient names in use before 1982 formed the starting baseline, leaving 1,273 unique names from 1982 to 2025.

### Character Trigram Surprisal

Each name was decomposed into character trigrams with boundary markers. For example, *Proscar* trigrams were *p, ^pr, pro, ros, osc, sca, car, ar*, *r$,* and the boundary markers (^ for start, $ for end) ensure that both prefix and suffix patterns contributed to the model.

Surprisal is a relative measure that has to be defined with respect to some reference set of names. A name is surprising or not with respect to some existing set of names, so defining this baseline set of names is critical. We built two separate baselines for computing surprisal, one for drug names and one for ordinary English. Thus we could compute whether a drug name’s trigrams were surprising compared to previously approved drug names, compared to ordinary English, or both.

The **drug name baseline** is computed separately for each name. For a name approved at time *t*, the trigram frequency model was built from all names that first appeared before *t* (names with the same date were not prior to each other). The drug name baseline also included the names already on the market or on file when the dataset began, most of which had no approval date (an augmented baseline).

We augmented the baseline because every new name competes with all names on the market. Without these additional names, NME names approved in 1985 would have been compared to 28 or fewer prior names. The NME baseline began with the 1,187 Orange Book trade names of NDA products approved before 1985 (1,030 of them undated because they were approved before 1982), and every later Orange Book NDA trade name and NME name entered at its first appearance (4,126 names in all). The other baselines began with the names in use before 1982 (Orange Book and generic) or filed before 1980 (USPTO). Augmenting names were never scored, and a name already in the baseline was not scored again (Supplementary Methods). We also did a sensitivity test using the unaugmented baseline (Supplementary Table S4).

If a trigram never appeared in the baseline, it would receive a probability of zero, and its surprisal (the negative logarithm of zero) would be infinite. To avoid this, we added a small constant (α = 0.1) to the count of every trigram and reserved one extra category (an “unknown bucket”) for all trigrams not yet observed. The probability of trigram *g* is

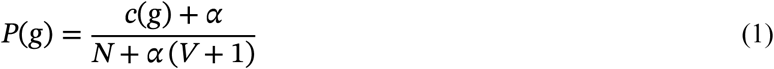

where *c*(*g*) is the count of *g* in the baseline, *N* is the total number of trigrams in the baseline, and *V* is the number of distinct trigrams. In sensitivity analyses, the direction and statistical significance of the trend did not change across values of alpha from 0.01 to 1.0, although its magnitude did (Supplementary Table S3).

Because *N* and *V* both grow as the number of baseline names grows, we checked whether baseline growth could produce the trend, by shuffling the approval dates among the scored names (5,000 shuffled datasets) and by rescoring every name against comparison sets that stayed the same for every year (Supplementary Methods).

The **English baseline (fixed)** is a trigram model of the 50,000 most common English words in the *wordfreq* library (version 3.1.1) [18], weighted by word frequency (counts per million words, at least 1). We computed two measures (drug name baseline and English baseline) for each name.

**Mean surprisal** measures how unexpected a name’s letter trigrams were, given the names (or words) in the baseline. Letter trigrams that have appeared often in the baseline data have low surprisal scores. Trigrams that appear rarely or never in the baseline data have high surprisal scores. A name with common trigrams (such as *pro* and *car* in *Proscar*) has low surprisal, whereas a name built from rare trigrams (such as *sdu* in *Jesduvroq*) has high surprisal. For a name with *k* trigrams *g*_1_ … *g_k_*, mean surprisal is

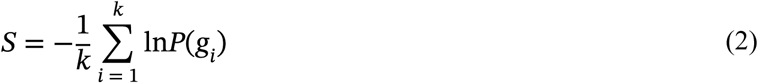

which is the average negative natural log of the smoothed trigram probabilities. We used natural logarithms, so surprisal is expressed in nats (the natural-logarithm unit of information). Higher values mean less predictable letter sequences. A rise of 1 nat in mean surprisal means that a name’s trigrams were, on average, about one-third as common in the baseline, and a rise of 1.5 nats means about one-fourth to one-fifth as common.

The **novel fraction** is the proportion of a name’s trigrams that never appeared in the baseline. With the drug name baseline, this is the proportion of trigrams never seen in any earlier name (the *drug baseline novel fraction*). With the English baseline, it is the proportion of trigrams that do not occur in any of the 50,000 English words (the *English novel fraction*). We also counted, by decade, the share of names containing at least one such trigram and the share containing a *q* not followed by *u*. To show the extremes, we listed the 12 NME names approved in 1985–1994 with the lowest English-baseline surprisal and the 12 approved in 2015–2024 with the highest (Table 1).

**Table 1.**
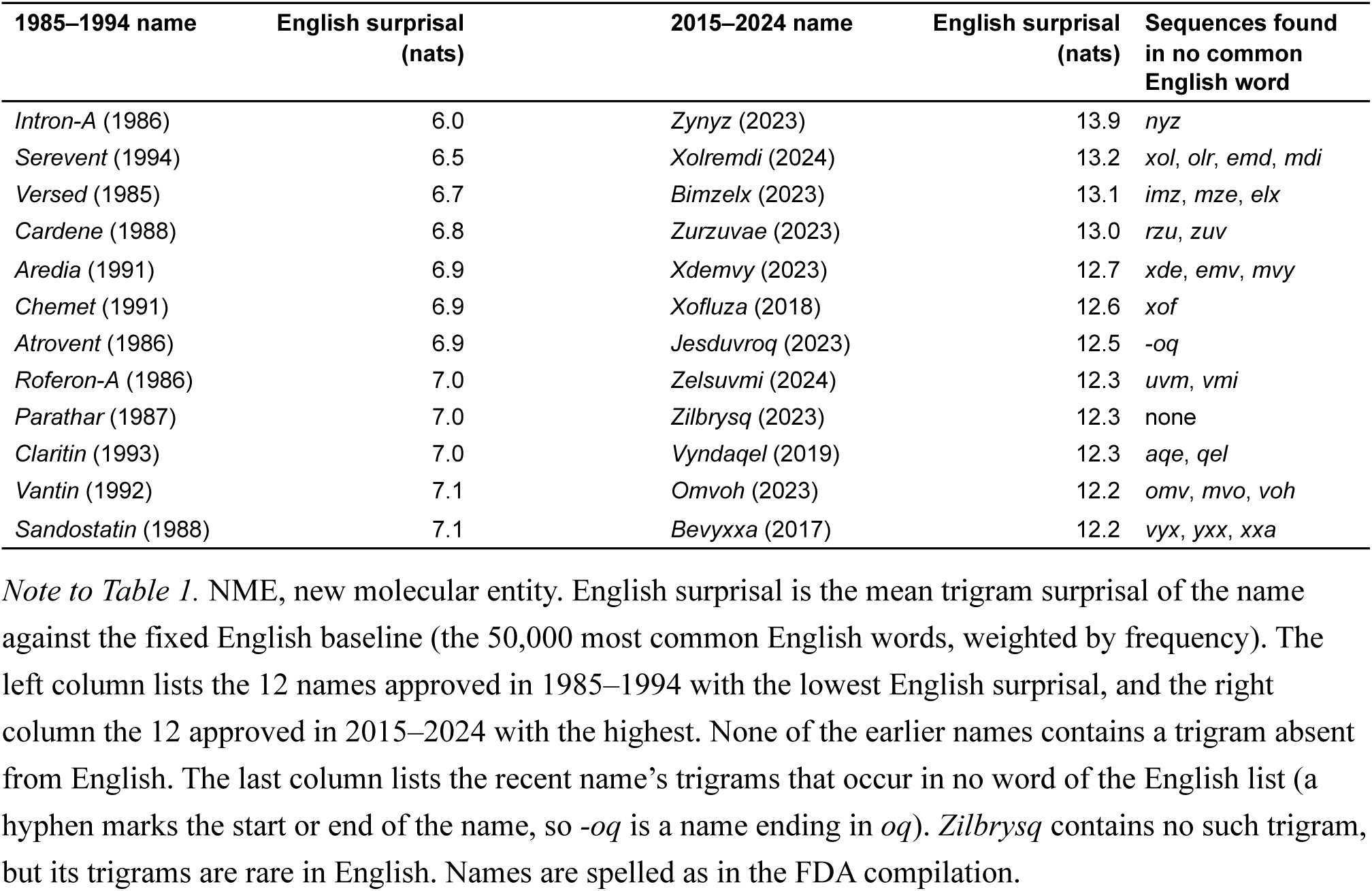
The most English-like NME names of 1985–1994 and the least English-like NME names of 2015–2024.

### Nearest-Neighbor Similarity and Namespace Density

To measure how crowded the namespace was, we computed the normalized Levenshtein similarity between each new name *a* and every previously approved name *b* (augmented baseline names included),

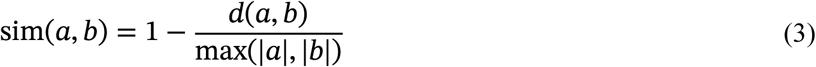

where *d*(*a*, *b*) is the edit distance and |*a*| is the number of characters in *a*, counting the space between the words of a multi-word name. Edit distance is the smallest number of single-letter insertions, deletions, or substitutions needed to turn one name into another (a similarity of 1 means identical spelling). For example, *Faslodex* and *Casodex* differ by two single-letter edits, and the longer name has eight letters, so their similarity is 1 − 2/8 = 0.75. For each name we took the mean similarity of its *N* closest neighbors (where *N* = 1, 3, 5, 10). We also counted how many prior names fell within fixed edit distances of 1, 2, and 3 (a measure sometimes referred to as neighborhood density).

Normalized Levenshtein similarity is analogous to, but simpler than, the similarity measures used internally by the FDA (e.g., the Phonetic and Orthographic Computer Analysis [POCA] system’s BI-SIM orthographic and ALINE phonetic-alignment algorithms [17]). Edit distance primarily measures spelling (look-alike) similarity, although it is also correlated with sound-alike similarity. It does not explicitly measure phonological/phonetic similarity. Greater crowding of the name space would tend to raise nearest-neighbor similarity over time, and active distancing would lower it.

### Statistical Analysis

We used linear regression to model trigram surprisal as a function of approval year and name length (in number of characters),

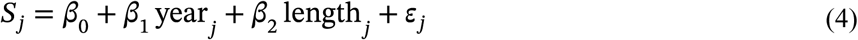

where *S_j_* is the mean surprisal of name *j*. We used robust standard errors to keep the confidence intervals correct even when the variability of scores differs from year to year. In a secondary model, we replaced approval year with the natural log of one plus the cumulative number of unique names approved before the current name. This model tested whether prior lexicon size predicted surprisal as well as calendar time did.

## Results

### What Recent Names Look Like

Table 1 sets the most English-like NME names of 1985–1994 beside the least English-like names of 2015– 2024. None of the earlier names contains a trigram absent from English. Eleven of the 12 recent names contain at least one, and *Xolremdi* contains four. Across all NME approvals, the share of names containing a letter sequence found in no common English word rose from 7% in the 1980s and 8% in the 1990s to 25% in the 2010s and 50% in the 2020s (Supplementary Table S8). No NME name contained a *q* not followed by *u* before *Pristiq* (2008). Since then 37 have, including *Kisqali*, *Nubeqa*, *Qelbree*, and *Jesduvroq*. Orange Book trade names showed the same rise (10% to 47% of names with a sequence absent from English), USPTO trademarks a smaller one (8% to 23%), and generic names none (16% to 18%).

Ranked against earlier drug names instead of English, the most surprising names were *Jeuveau* (2019, 10.7 nats, 44% novel trigrams), which borrows French orthography, *Jesduvroq* (2023, 10.3 nats), which chains three novel trigrams including *sdu*, and *Omvoh* (2023, 10.1 nats), which squeezes three novel English trigrams into five letters (Supplementary Table S1). The average NME name approved in the 1980s scored 7.21 nats. Low-surprisal names read like ordinary words (*Vantin*, *Lodine*, *Prandin*, *Detrol*). Several strategies recur, including *q* without *u*, unusual initial clusters (*Tzield*, *Ztalmy*), *x* and *z* (*Xolremdi*, *Xdemvy*), and unusual vowel sequences (*Jeuveau*, *Zurzuvae*). We offer these patterns as illustrations, without evaluation.

### Descriptive Trends

Table 2 gives trigram surprisal by decade across all four datasets (see also Figures 1 and 3). Mean trigram surprisal rose in every decade in all three brand-name datasets.

**Table 2.**
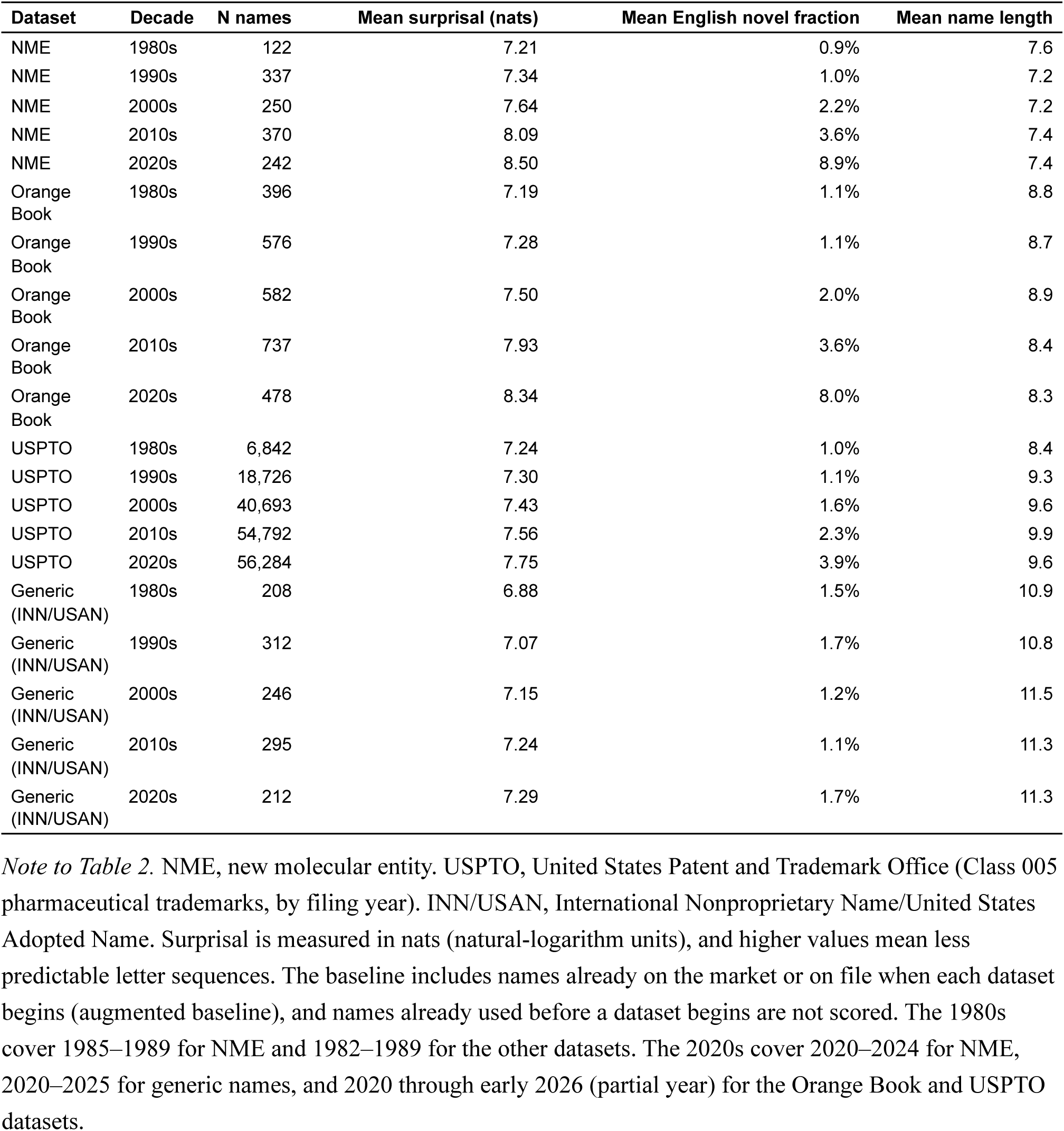
Mean drug baseline character trigram surprisal by decade across four datasets.

| Dataset | Decade | N names | Mean surprisal (nats) | Mean English novel fraction | Mean name length |
| --- | --- | --- | --- | --- | --- |
| NME | 1980s | 122 | 7.21 | 0.9% | 7.6 |
| NME | 1990s | 337 | 7.34 | 1.0% | 7.2 |
| NME | 2000s | 250 | 7.64 | 2.2% | 7.2 |
| NME | 2010s | 370 | 8.09 | 3.6% | 7.4 |
| NME | 2020s | 242 | 8.50 | 8.9% | 7.4 |
| Orange Book | 1980s | 396 | 7.19 | 1.1% | 8.8 |
| Orange Book | 1990s | 576 | 7.28 | 1.1% | 8.7 |
| Orange Book | 2000s | 582 | 7.50 | 2.0% | 8.9 |
| Orange Book | 2010s | 737 | 7.93 | 3.6% | 8.4 |
| Orange Book | 2020s | 478 | 8.34 | 8.0% | 8.3 |
| USPTO | 1980s | 6,842 | 7.24 | 1.0% | 8.4 |
| USPTO | 1990s | 18,726 | 7.30 | 1.1% | 9.3 |
| USPTO | 2000s | 40,693 | 7.43 | 1.6% | 9.6 |
| USPTO | 2010s | 54,792 | 7.56 | 2.3% | 9.9 |
| USPTO | 2020s | 56,284 | 7.75 | 3.9% | 9.6 |
| Generic (INN/USAN) | 1980s | 208 | 6.88 | 1.5% | 10.9 |
| Generic (INN/USAN) | 1990s | 312 | 7.07 | 1.7% | 10.8 |
| Generic (INN/USAN) | 2000s | 246 | 7.15 | 1.2% | 11.5 |
| Generic (INN/USAN) | 2010s | 295 | 7.24 | 1.1% | 11.3 |
| Generic (INN/USAN) | 2020s | 212 | 7.29 | 1.7% | 11.3 |

*Note to Table 2.* NME, new molecular entity. USPTO, United States Patent and Trademark Office (Class 005 pharmaceutical trademarks, by filing year). INN/USAN, International Nonproprietary Name/United States Adopted Name. Surprisal is measured in nats (natural-logarithm units), and higher values mean less predictable letter sequences. The baseline includes names already on the market or on file when each dataset begins (augmented baseline), and names already used before a dataset begins are not scored. The 1980s cover 1985–1989 for NME and 1982–1989 for the other datasets. The 2020s cover 2020–2024 for NME, 2020–2025 for generic names, and 2020 through early 2026 (partial year) for the Orange Book and USPTO datasets.

**Figure 1.**
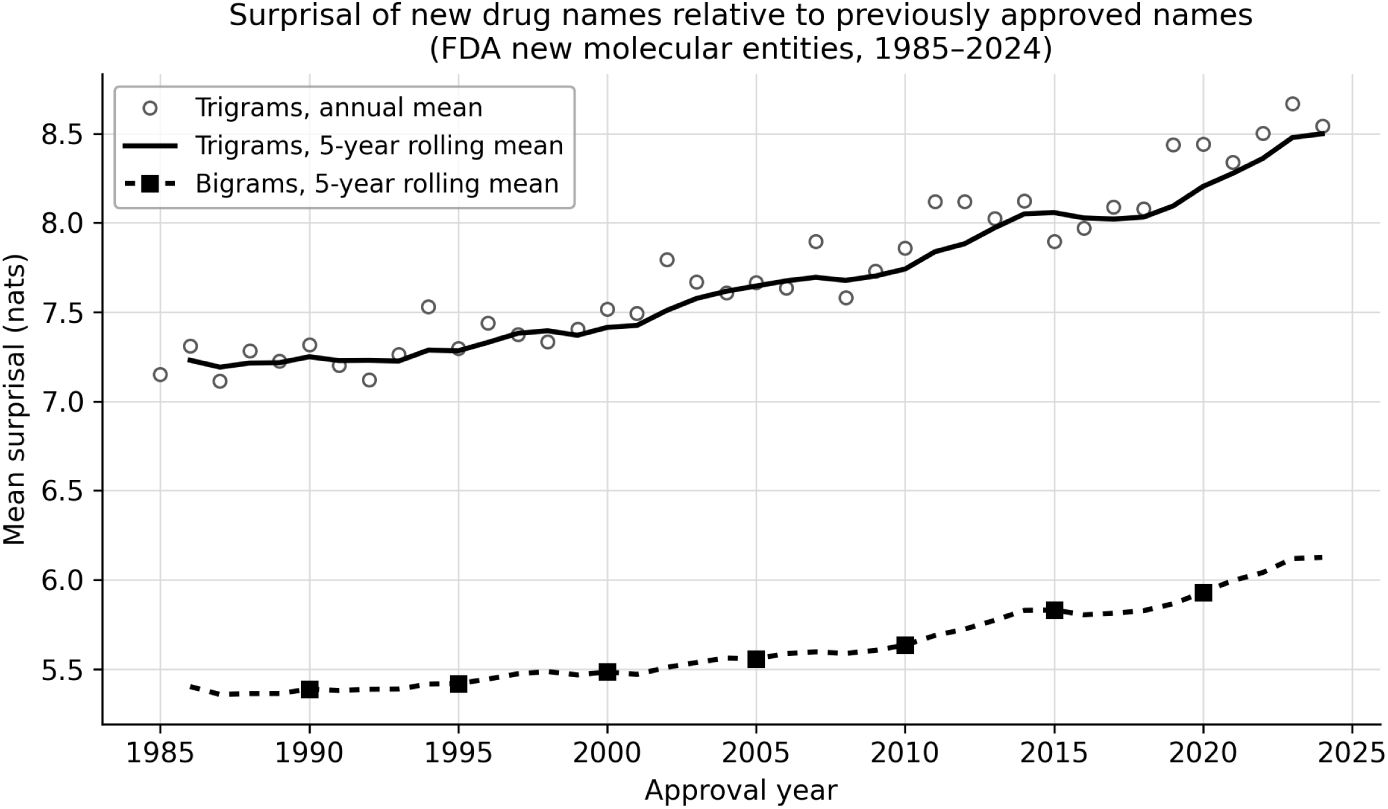
Mean drug baseline surprisal by approval year (NME dataset, 1985–2024). Prior names include the 1,187 Orange Book trade names approved before 1985 (augmented baseline) and every later Orange Book trade name and NME name from its first appearance. Open circles show the annual mean trigram surprisal of all names approved that year. The solid line shows the 5-year rolling mean of trigram surprisal. The dashed line with filled squares shows the 5-year rolling mean of bigram (two-letter sequence) surprisal, included for comparison. Surprisal is in nats, and higher values indicate names with less predictable character sequences relative to previously approved names.

Surprisal in the NME dataset increased from 7.21 nats in the 1980s to 8.50 nats in the 2020s, an 18% increase. The Orange Book followed a similar pattern (7.19 to 8.34 nats, 16% increase). The USPTO data showed the same trend, although the increase there (7.24 to 7.75 nats, 7% increase) was smaller, perhaps because Class 005 includes dietary supplements and cosmeceuticals whose conventional English-word marks (e.g., “HEALTHPLUS”) dilute the trend. Generic (INN/USAN) names rose much more slowly, from 6.88 to 7.29 nats, a 6% increase (Figure 3). Restricted to single-word names, every proprietary-name year coefficient was as large or larger (Supplementary Table S6).

The drug baseline novel fraction was 0.07 in both the late 1980s and the 1990s, and then more than doubled, to 0.15 in the 2020s (Supplementary Figure S1). Thus, about one in seven trigrams in recent names is new.

### Regression Results

Approval year (filing year for USPTO) was positively associated with drug baseline trigram surprisal in all four datasets (Table 3). The per-year coefficient was largest in the NME dataset (b = 0.038) and smallest in the generic dataset (b = 0.011). Over the 39-year NME period, this coefficient implies a cumulative increase of approximately 1.5 nats. In other words, the letter sequences in a 2024 name were, on average, about one-fourth to one-fifth as common in earlier names as those in a 1985 name.

**Table 3.**
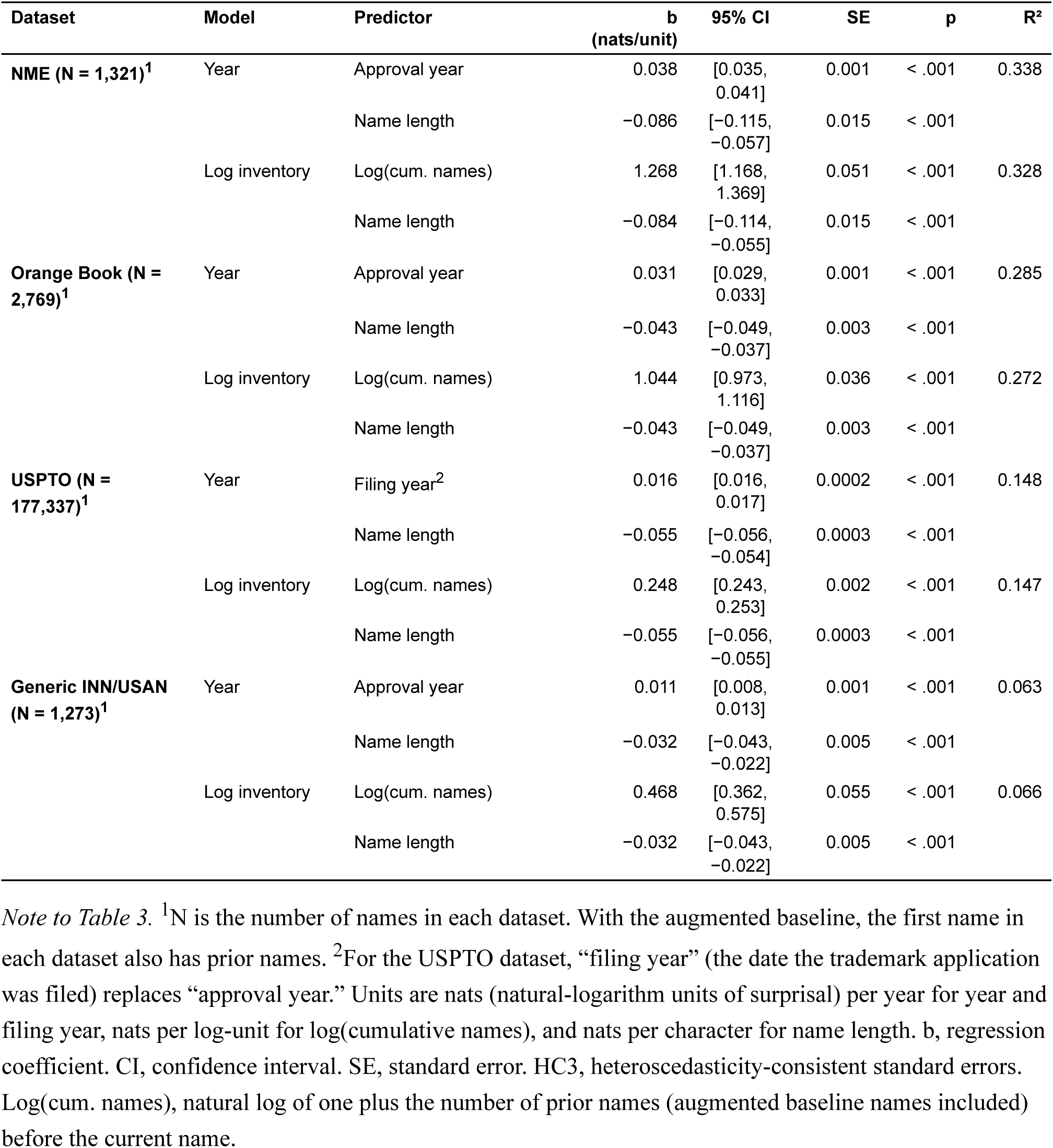
Predictors of drug baseline trigram surprisal across four datasets (ordinary least squares regression with HC3 robust standard errors).

| Dataset | Model | Predictor | b<br>(nats/unit) | 95% CI | SE | p | R<sup>2</sup> |
| --- | --- | --- | --- | --- | --- | --- | --- |
| **NME (N = 1,321)<sup>1</sup>** | Year | Approval year | 0.038 | [0.035, 0.041] | 0.001 | < .001 | 0.338 |
|  |  | Name length | -0.086 | [-0.115, -0.057] | 0.015 | < .001 |  |
|  | Log inventory | Log(cum. names) | 1.268 | [1.168, 1.369] | 0.051 | < .001 | 0.328 |
|  |  | Name length | -0.084 | [-0.114, -0.055] | 0.015 | < .001 |  |
| **Orange Book (N = 2,769)<sup>1</sup>** | Year | Approval year | 0.031 | [0.029, 0.033] | 0.001 | < .001 | 0.285 |
|  |  | Name length | -0.043 | [-0.049, -0.037] | 0.003 | < .001 |  |
|  | Log inventory | Log(cum. names) | 1.044 | [0.973, 1.116] | 0.036 | < .001 | 0.272 |
|  |  | Name length | -0.043 | [-0.049, -0.037] | 0.003 | < .001 |  |
| **USPTO (N = 177,337)<sup>1</sup>** | Year | Filing year<sup>2</sup> | 0.016 | [0.016, 0.017] | 0.0002 | < .001 | 0.148 |
|  |  | Name length | -0.055 | [-0.056, -0.054] | 0.0003 | < .001 |  |
|  | Log inventory | Log(cum. names) | 0.248 | [0.243, 0.253] | 0.002 | < .001 | 0.147 |
|  |  | Name length | -0.055 | [-0.056, -0.055] | 0.0003 | < .001 |  |
| **Generic INN/USAN (N = 1,273)<sup>1</sup>** | Year | Approval year | 0.011 | [0.008, 0.013] | 0.001 | < .001 | 0.063 |
|  |  | Name length | -0.032 | [-0.043, -0.022] | 0.005 | < .001 |  |
|  | Log inventory | Log(cum. names) | 0.468 | [0.362, 0.575] | 0.055 | < .001 | 0.066 |
|  |  | Name length | -0.032 | [-0.043, -0.022] | 0.005 | < .001 |  |

*Note to Table 3.* <sup>1</sup>N is the number of names in each dataset. With the augmented baseline, the first name in each dataset also has prior names. <sup>2</sup>For the USPTO dataset, “filing year” (the date the trademark application was filed) replaces “approval year.” Units are nats (natural-logarithm units of surprisal) per year for year and filing year, nats per log-unit for log(cumulative names), and nats per character for name length. b, regression coefficient. CI, confidence interval. SE, standard error. HC3, heteroscedasticity-consistent standard errors. Log(cum. names), natural log of one plus the number of prior names (augmented baseline names included) before the current name.

Baseline growth did not produce this trend. With the approval dates shuffled at random, the NME year coefficient averaged −0.010 (95% of the 5,000 shuffled datasets between −0.014 and −0.007), and none of them reached 0.038. A growing baseline lowers surprisal, so the observed trend is conservative (i.e., surprisal increases in spite of the growing lexicon size). The other datasets, and comparison sets that stayed the same for every year, gave the same result (Supplementary Table S5).

The secondary model, using log(cumulative unique names before), fit about as well in all datasets, but inventory size and calendar time are too highly correlated to separate. Year coefficients were similar with the unaugmented baseline (Supplementary Table S4).

Longer names had lower mean surprisal in all four datasets (b = −0.032 to −0.086, p < .001).

The year effect was also strong in the English-baseline model (b = 0.049, 95% CI [0.044, 0.054], p < .001, R² = 0.243). Relative to English, mean NME surprisal rose from 8.22 nats in the 1980s to 9.97 nats in the 2020s.

The English novel fraction rose from less than 1% in the 1980s to approximately 9% on average in the 2020s, reaching 13% by 2024 (a roughly tenfold increase, Supplementary Figure S2). Recent names such as *Omvoh* (novel English trigrams *omv*, *mvo*, *voh*), *Zurzuvae* (*rzu*, *zuv*), and *Kisqali* (*sqa*, *qal*) contain character sequences with no precedent in the 50,000 most common English words.

### Nearest-Neighbor Similarity and Namespace Density

Nearest-neighbor similarity declined from the early 1990s onward (Figure 2). The mean similarity between a new name and its closest prior neighbor stayed between 0.61 and 0.62 from 1987 through 1992 and then fell to 0.59 by 2005 and 0.54 by 2024 (top-1 neighbor, 5-year rolling mean). In letters, a new name in the 1990s could be turned into its closest prior name with about three single-letter changes, and by the 2020s it took about three and a half, in names about seven or eight letters long. Mean top-10 similarity peaked at 0.52 in 1992 and fell to 0.47 by 2024. During the same period the number of prior names grew more than threefold (from 1,194 to 4,034 names).

**Figure 2.**
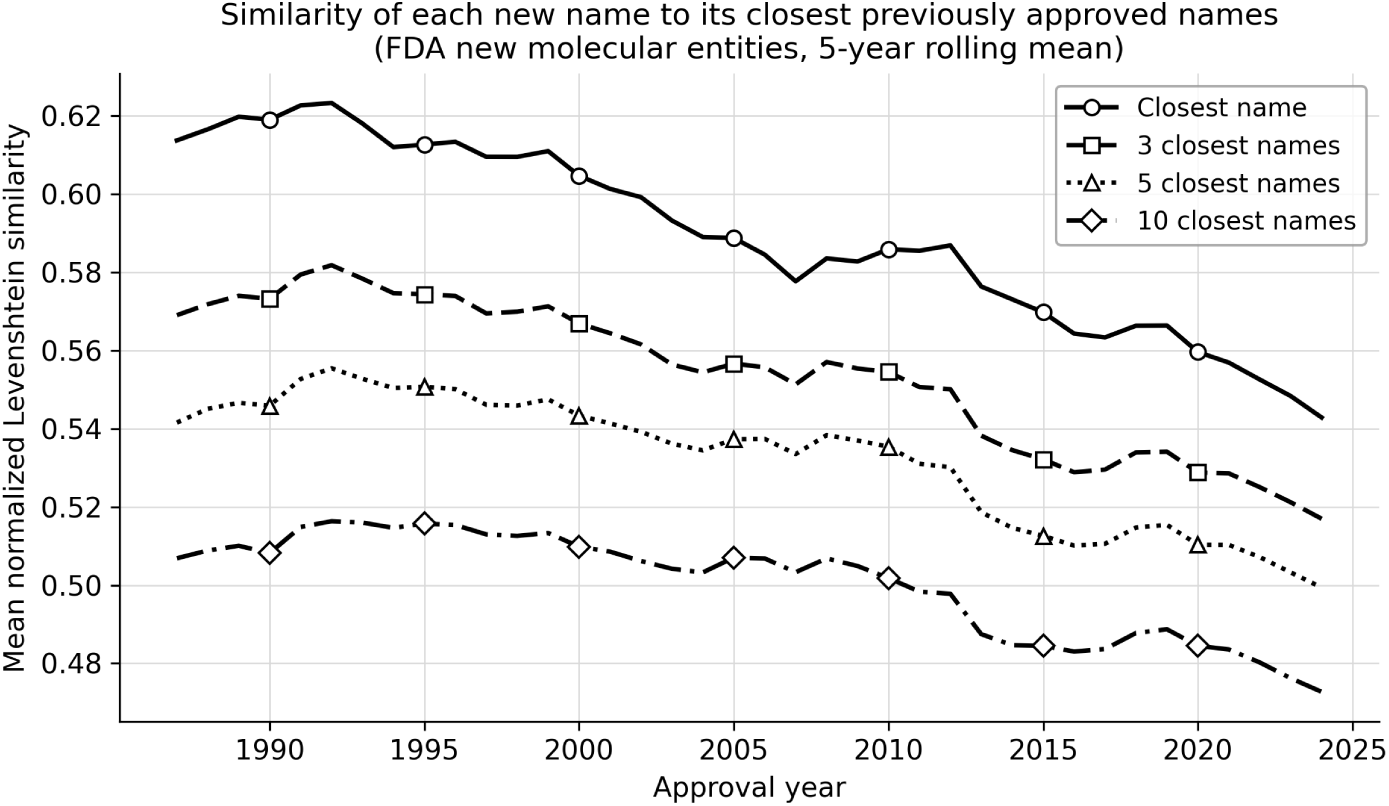
Mean nearest-neighbor similarity (normalized Levenshtein similarity, where 1 means identical spelling) by approval year, averaged over the closest 1, 3, 5, and 10 prior names (NME dataset, 5-year rolling mean). Prior names include the 1,187 Orange Book trade names approved before 1985 (augmented baseline) and every later Orange Book trade name and NME name from its first appearance. Line styles and marker shapes distinguish the four series. Similarity was highest in the early 1990s and declined thereafter, although the lexicon of prior names more than tripled.

The neighborhood density analysis also showed a decline, after an earlier rise (Supplementary Figure S5). Among Orange Book brand names, the mean number of prior names within edit distance 3 was 2.84 in the 1980s, climbed to 3.97 in the 1990s and 3.71 in the 2000s, and fell back to 2.13 in the 2020s. A regression on approval year and name length gave a negative year coefficient (Orange Book, b = −0.045, p < .001, R² = 0.174), and a squared year term improved the fit slightly (b = −0.0034, p < .001, R² = 0.185), with a peak in 1998.

There was no trend in generic name neighborhood density. Mean count within distance 3 was 0.64 in the 1980s, 0.60 in the 2010s, and 0.58 in the 2020s, and the year coefficient was not significant (b = 0.000, p = .94, R² = 0.03). Generics had fewer close neighbors than brands (1.38 fewer, adjusted for name length), and two-thirds of this gap (63%) came from the larger brand lexicon (Supplementary Methods).

Despite progress in coining new names that are increasingly distant from existing names, some very similar names were still approved. Among post-2000 NME approvals, 33 had a prior FDA-approved name with similarity of 0.70 or more (Supplementary Table S7). In 19 of these pairs the earlier product is now listed as discontinued, and in 12 both products are still marketed. Among pairs in which both products are still marketed, the closest were *Besivance* and *Kepivance* (similarity = 0.78), *Faslodex* and *Casodex* (0.75), and *Septocaine* and *Sensorcaine* (0.73), both injectable local anesthetics. These scores are objective measures of string similarity, not judgments of clinical confusion risk (which also depends on dose, form, route, and setting).

### Replications and Generic Comparison

All major findings were replicated in the Orange Book, a larger and more diverse sample, although not an independent one (most NME drugs also appear in the Orange Book, and the NME baseline includes Orange Book trade names). Its year coefficient was smaller (b = 0.031), as was its model fit (R² = 0.29 vs. 0.34). The English novel fraction increased from 1.1% to 8.0%, and the nearest-neighbor analysis showed the same decline (top-1 similarity peaked at 0.62 in 1988 and declined to 0.56 by the 2020s).

The USPTO analysis replicated the core finding (Table 2, Supplementary Figure S3). Filing year predicted surprisal (b = 0.016, p < .001, R² = 0.15). The English novel fraction also rose over the period, from 1.0% to 3.9% (Supplementary Figure S4). Unlike FDA names, USPTO marks did not become less similar to their nearest neighbors. Similarity to the nearest neighbor rose from 0.66 in 1980–1984 to 0.70 in 2000–2004 and stayed at 0.70 through 2026, while the number of earlier marks quadrupled. The mean number of earlier marks within edit distance 3 rose from 23 to 170 as the register grew.

Generic (INN/USAN) names showed a trend that was significant but weaker than that for brand names (Figure 3). The year coefficient was about one-third the magnitude of the Orange Book brand value (b = 0.011 vs. 0.031), and the model explained less variance (R² = 0.063 vs. 0.29).

**Figure 3.**
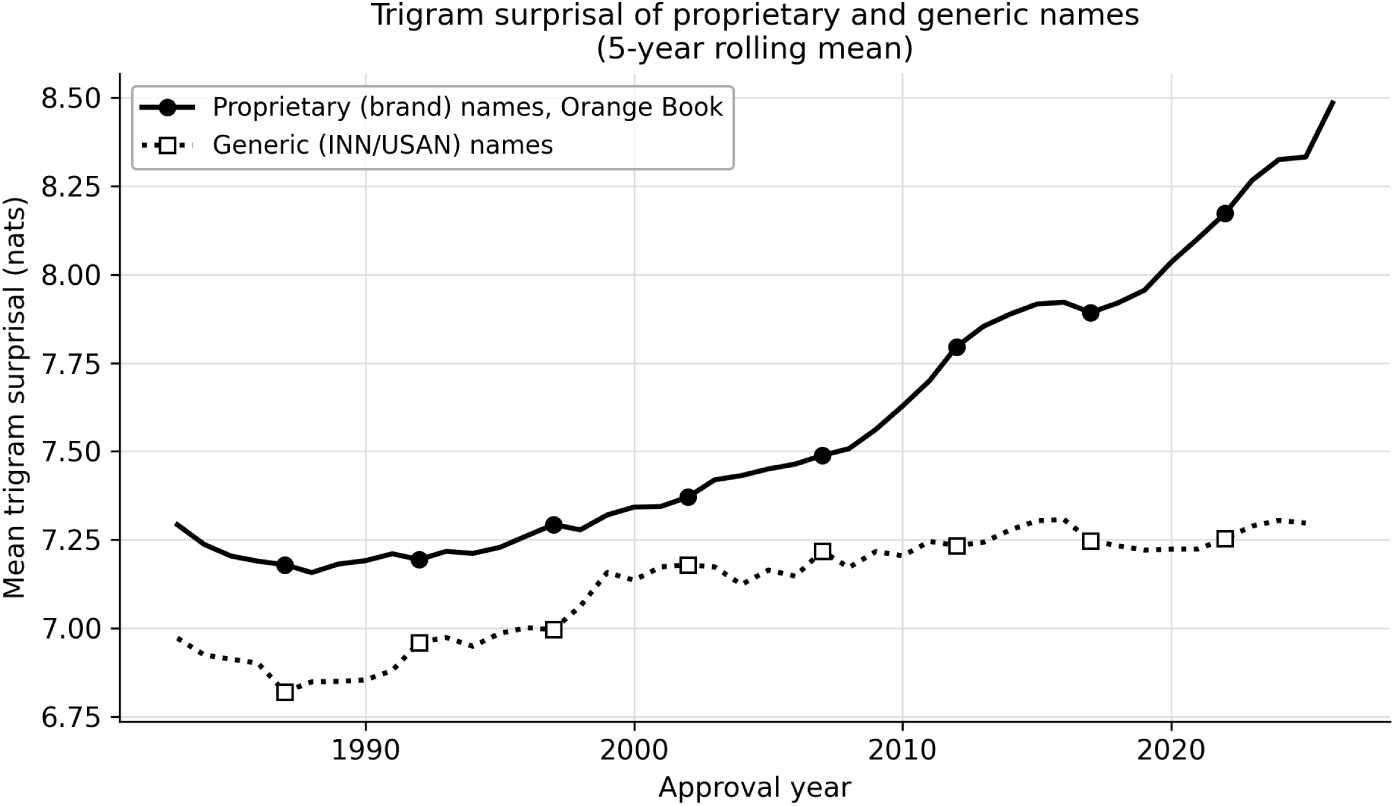
Mean drug baseline character trigram surprisal by approval year for proprietary names (Orange Book brand names, solid line with filled circles) and generic (INN/USAN) names (dotted line with open squares), 1982–2026 (5-year rolling means, with 2026 a partial year). Prior names include names approved before 1982 (augmented baseline). Names already used before 1982 are not scored. Proprietary names show a steeper upward trend, consistent with greater degrees of freedom for orthographic innovation in the absence of INN stem constraints.

The weaker trend might be explained by the enforced stem system in INNs. The INN stem system requires pharmacologically related drugs to share stems, and these shared stems constantly recycle the same trigrams and reduce surprisal. Proprietary names, in contrast, can use the entire orthographic space.

Across decades, the generic English novel fraction was low and essentially flat (1.1–1.7%). The pattern likely reflects the Latin and Greek morphemes in generic names, whose character patterns overlap substantially with English.

## Discussion

Across four decades of FDA approvals, drug names became more orthographically distinctive. Half of the NME names approved in the 2020s contain a letter sequence found in no common English word, and surprisal relative to prior drug names went from 7.21 to 8.50 nats (an 18% increase). Although the lexicon more than tripled in size, nearest-neighbor similarity declined from the early 1990s onward, and edit-distance neighborhood counts among brand names fell after an earlier rise.

The USPTO replication covered 177,337 trademarks (including proposed-but-never-approved names) and showed that the trend extends beyond FDA-approved names, although it cannot rule out changes in FDA selection. Generic names showed a significant but weaker version of the trend.

The idea that orthographic distinctiveness reduces the risk of confusion is based on experimental evidence. Orthographic similarity increases misidentification, false recognition, and recall errors [10–12], so lower similarity to existing names might, all else being equal, reduce confusion. We did not measure confusion.

For FDA names, nearest-neighbor similarity fell even as the number of names in the database grew. This is consistent with one or more active distancing forces (FDA name review, ISMP advocacy, industry self-policing, or shifts in naming style) working against the tendency toward crowding. FDA review of proposed names for confusability began around 1990 in CDER’s Labeling and Nomenclature Committee [19, 20], about when the decline began, and passed to dedicated medication error staff in 1999 [20]. The timing is consistent with a contribution from FDA review but does not single out any one causal factor.

Computerized screening might also have contributed. The FDA’s POCA system was built on the BI-SIM orthographic and ALINE phonetic algorithms [17]. When FDA staff analyzed confused name pairs from the ISMP list, 99% shared the same first letter, a three-or-more letter string, or similar length [21].

Longer names had lower surprisal, including among single-word names. A name built entirely from unfamiliar trigrams would be hard to read and pronounce, so a longer name is more likely to include some familiar letter sequences. These appear to come from earlier drug names (device names such as *HandiHaler*, INN stems, common prefixes and suffixes), because against the English baseline length had little or no association with surprisal. NME name length changed little (between 7.2 and 7.6 characters), so it does not account for the rise in surprisal.

Orthographic distinctiveness matters most in tasks involving visual perception (e-prescribing, printed labels, electronic health record lists, pharmacy shelving). It might matter less in tasks involving auditory perception, where phonological similarity is more important. Unfamiliar spelling might also have detrimental effects on populations with limited English proficiency, low health literacy, or cognitive impairment.

### Limitations

One concern involves the generic analysis, which treats each name as independent. The INN stem system introduces structured dependence (all 73 *-tinib* names share the trigrams tin, ini, nib, ib$, and b$$), and one direction for future work is to decompose surprisal into stem and non-stem components. The dynamic baseline also treats all prior names equally regardless of market status, because a withdrawn name still occupies namespace.

The augmented baseline name lists are a lower bound on the names in use when each dataset begins. They omit products withdrawn before the Orange Book began in 1980, and the pre-1980 USPTO records are mostly registered marks, because applications abandoned before 1980 are nearly absent from the USPTO file. Biologics licensed before 1985 and biosimilars are absent from every baseline. Names approved in the same year might resemble one another in ways the model does not capture. When we grouped the standard errors by approval year (cluster-robust standard errors), the confidence intervals widened in all four datasets (by factors of 1.08 to 1.81, and 5.86 for the trademark data), but every year trend remained significant (p < .001).

Trigram surprisal captures orthographic (but not phonological) similarity, so future work could incorporate phoneme-level models. For the nearest-neighbor analysis we used a simple global edit distance. Metrics of similarity weighted by visual or phonological confusability would come closer to real error conditions [3].

We documented time trends. But precisely what caused these time trends is unclear. The contributions of regulation, marketing, or trademark congestion remain confounded. Nor did the present study test whether increased distinctiveness reduced error rates.

### Implications

Our results are consistent with a conscious effort by industry and regulators to make drug names less confusable, and with that effort succeeding, at least in producing novel spellings and distance from existing names. Unusual spelling has benefits and costs. A name with unusual character sequences is less likely to resemble existing names. This might lower the chance of visual confusion. But such a name might be harder to read, pronounce, spell, and remember, and this could cause errors elsewhere in the medication-use process.

Text enhancements such as Tall Man (i.e., mixed case) lettering have produced mixed results [22, 23]. Building distinctiveness into the spelling of a name when it is coined might prove more durable than typographic fixes applied after the fact (although the best trade-off between distinctiveness and familiarity is unknown [24]).

For regulators, these findings are consistent with the intuition and intention that the name review process, taken as a whole, contributed to increased distinctiveness. For industry, our findings highlight the value of computational tools that evaluate a candidate name’s distinctiveness before submission. Such tools belong in a multi-criteria screening process (alongside phonetic assessment, handwriting simulation, and human factors testing).

The nearest-neighbor analysis flagged some highly similar pairs of names as well (e.g., *Besivance*/*Kepivance*, *Faslodex*/*Casodex*, *Septocaine*/*Sensorcaine*). Because both names in each of these pairs are still marketed, they could warrant continued pharmacovigilance. If the trend of increasing distinctiveness continues, interested parties should consider the need for a higher standard of evidence that increasingly unfamiliar names do not introduce new forms of error, even if they are less similar to existing names.

## Conclusion

Over 40 years of FDA approvals, newly approved brand drug names became more orthographically distinctive, both relative to previously approved names and relative to ordinary English. By the 2020s, half of new names contained a letter sequence found in no common English word, a trend also seen in USPTO pharmaceutical trademarks. The trend was present but weaker in generic names. Since the 1990s, new names have had fewer and more distant nearest neighbors, a pattern consistent with conscious efforts to decrease similarity and confusability, although the present data cannot identify specific causes. Increasingly unusual spellings of drug names should not necessarily be equated with a reduced rate of name confusion errors, because unfamiliar letter patterns might be harder to read, spell, and remember.

## Author Contributions and Use of Artificial Intelligence

B.L. conceived of the project, identified the data sources, developed the research questions and hypotheses, planned the analyses, identified key articles from the previous literature, interpreted the results, and revised and finalized the manuscript. The author used AI coding assistants (Claude Code from Anthropic and Codex from OpenAI) to write and run most of the analysis code under the author’s supervision. These systems assisted in drafting some sections of the initial manuscript, including plain-language explanations of statistical terms, and provided language revision of later drafts.

The author reviewed all of the code output, checked every reported number against the analysis results, revised the AI-drafted text, and approved the final version. All data are publicly available, and no confidential material was shared with an AI system. The author takes full responsibility for the content of the article.

## Data Availability

All data analyzed in this study are publicly available. Proprietary drug names were obtained from the FDA Center for Drug Evaluation and Research Compilation of New Molecular Entity (NME) Drug and New Biologic Approvals, 1985–2024 (https://www.fda.gov/drugs/drug-approvals-and-databases/compilation-cder-new-molecular-entity-nme-drug-and-new-biologic-approvals). Trade names and generic ingredient names were obtained from the FDA Orange Book data files, Products file (https://www.fda.gov/drugs/drug-approvals-and-databases/orange-book-data-files). Both FDA files were downloaded in February 2026. Pharmaceutical trademarks (International Class 005) were extracted from the US Patent and Trademark Office Trademark Full-Text XML Data, Applications, product TRTYRAP, annual back file, 2025 edition (files dated December 31, 2025, downloaded February 2026), which contains applications filed through February 9, 2026 (https://data.uspto.gov/bulkdata/datasets/trtyrap). English word frequencies were obtained from the wordfreq library, version 3.1.1 (Speer R., 2023, https://pypi.org/project/wordfreq/3.1.1/, archived versions at https://doi.org/10.5281/zenodo.595730).

The analysis code, the USPTO extraction code, the FDA source files as downloaded, the extracted USPTO Class 005 records, and the scored name-level datasets for all four datasets (primary and unaugmented baselines), together with the lists of Orange Book entries excluded as not being trade names, are deposited in Zenodo (https://doi.org/10.5281/zenodo.23196771).

## Online Supplementary Material

### Supplementary Methods

#### New Molecular Entity (NME) Compilation

We excluded placeholder entries. Three entries were scored as a single trade name. Two cells each listed two trade names (*Lotemax* with *Alrex*, and *Maxalt* with *Maxalt-MLT*), and we scored the first name of each. The second names are Orange Book trade names and entered the baseline at their Orange Book dates. A third entry was a descriptive phrase followed by its trade name (*Skin Exposure Reduction Paste Against Chemical Warfare Agents (SERPACWA)*), and we scored *SERPACWA*. Scored as written, this 62-letter entry was by far the longest NME name, and it alone made the name-length coefficient of the NME year model nonsignificant (b = −0.043, p = .38). Scored as *SERPACWA*, name length had a negative coefficient, as in the other datasets (Table 3). In the Orange Book, two trade names (PREMPHASE and PREMPRO) were followed by a parenthesized list of their component products, and we removed the list.

The compilation lists both new drug applications (NDAs) and biologics license applications (BLAs). Of the 1,321 scored names, 1,055 came from NDAs and 266 from BLAs. Biologics are not in the Orange Book, and none of the 266 BLA names is spelled like an Orange Book trade name, so each BLA name entered the NME baseline at its NME approval date. Of the NDA names, 61 have no Orange Book trade name with the same spelling (for example, withdrawn products such as *Seldane* and renamed products such as *Losec*). These names also entered at their NME approval dates. One NME name, *Imagent* (2002), had already been used for a different Orange Book product in 1993, so it was treated as a name already in use and was not scored.

#### Orange Book Entries That Are Not Trade Names

The trade-name field of the Orange Book also holds entries that are not trade names. We excluded three groups of them. Descriptive product titles were names whose words, after normalization, included in, and, w (from W/, meaning “with”), or container (e.g., *potassium chloride in dextrose and sodium chloride in plastic container*, *ancef in dextrose in plastic container*, *sulfamethoxazole and trimethoprim and phenazopyridine hydrochloride*, and *phenergan w codeine* from PHENERGAN W/ CODEINE). Group A held names with no coined trade name, which were either the established name of the product (the trade name was spelled like the ingredient field of an Orange Book product, e.g., *diltiazem hydrochloride*, *potassium iodide*, and *sterile water for injection*) or one of six reviewed descriptive titles that the other criteria missed (e.g., *skin exposure reduction paste against chemical warfare agents* and *leuprolide acetate for depot suspension*). Group B held names of the same form as the descriptive titles but written with “&” or with the word “with” (e.g., *Aleve-D Sinus & Cold* and *Advil Dual Action with Acetaminophen*). Names that join a trade name to a descriptor such as Kit, Autoinjector, or Preservative Free were kept. Excluded entries were not scored and were not added to any baseline, so they were not prior names in the nearest-neighbor and neighborhood-count analyses either.

The rule removed 395 of the 3,164 Orange Book names that would otherwise have been scored, which left 2,769. These were 221 descriptive titles, 165 in group A, and 9 in group B. The excluded entries were concentrated in the early years. They made up 148 of the 544 names first used in the 1980s (27%), 62 of 638 in the 1990s (10%), 56 of 638 in the 2000s (9%), 75 of 812 in the 2010s (9%), and 54 of 532 in the 2020s (10%). By decade from the 1980s to the 2020s, the descriptive titles numbered 114, 42, 20, 26, and 19, and the group A names numbered 33, 18, 34, 48, and 32. The same rule removed 168 of the 1,198 undated pre-1982 augmenting names (85 titles, 82 in group A, and 1 in group B), which left 1,030 Orange Book augmenting names. The NME baseline is built from the same Orange Book trade names, so the rule also removed 241 of the 1,428 NME augmenting names (which left 1,187) and 322 of the 1,940 Orange Book trade names that entered the NME baseline from 1985 on (which left 1,618). NME names, generic names, and USPTO marks were not changed, and the generic and USPTO results are the same with and without the rule. A few excluded titles join a trade name to a description (e.g., *Advil Cold and Sinus* and *Fiorinal W/Codeine*). We applied the rule as defined, with no exceptions.

#### Units of Analysis and Multi-Word Names

Each unique name was one observation in every analysis, so the sample sizes in Tables 1 and 2 are numbers of names, and the robust standard errors treat names as independent observations. Names were converted to lower case, trademark symbols were removed, the characters - / _ . (), : and + were replaced by spaces, and the name was split into words. A final dosage-form word (XR, ER, SR, CR, ODT, IR, DR, LA, XL, EC, MR, HR, IV, IM, or SC) was removed when the name had more than one word, and digits were removed. Trigrams were computed within each word, with boundary markers on each word, so no trigram spanned two words. A name’s surprisal and novel fraction were means over all trigrams of all its words, so each word was weighted by its number of trigrams, and a trigram that occurred twice in a name counted twice. Repeated trigrams were rare (4 of 12,393 NME trigrams, 0.3% of generic trigrams, 0.6% of Orange Book trigrams, and 1.7% of USPTO trigrams). After a name was scored, all of its trigrams were added to the baseline. Name length was the number of letters in all words of the name. For edit distances (nearest-neighbor similarity and neighborhood counts), each name was written as its words separated by single spaces. The separator characters listed above therefore became one space, and hyphenated and spaced forms of a name gave the same string. Spaces counted as characters both in the distance and in the length used to normalize it.

Multi-word names were more common in early Orange Book records. Their share fell from 26% of Orange Book names in the 1980s to 13% in the 2020s (NME 5.7% to 0.4%, USPTO 28% to 35%, generic 10.6% to 2.8%). We therefore repeated the year models on single-word names only, both against the full baseline and against a baseline built from single-word names only (Supplementary Table S6). Every proprietary-name year coefficient was as large or larger than with all names. For the Orange Book, single-word names rose from 7.26 nats in the 1980s to 8.45 nats in the 2020s, and R² was 0.29 with all names and 0.30 with single-word names. Nearest-neighbor similarity also declined among single-word names (NME top-1 similarity, b = −0.0023 per year, Orange Book b = −0.0019, both p < .001), as did Orange Book neighborhood counts (b = −0.066, p < .001). Generic neighborhood counts again showed no trend (p = .89). As a further check, we recomputed nearest-neighbor similarity and neighborhood counts from the first word of every name, for scored and prior names alike. Every trend kept its direction. NME closest-name similarity peaked at 0.63 in 1992 and fell to 0.55 by 2024, and the Orange Book count of prior names within distance 3 fell from 9.08 in the 1980s to 3.98 in the 2020s (b = −0.14 per year, p < .001).

#### United States Patent and Trademark Office (USPTO) Pharmaceutical Trademark Filter

Marks were kept if their goods and services description contained pharmaceutical-relevant keywords (e.g., *pharmaceutical*, *prescription*, *tablet*, *vaccine*), and veterinary-only marks were excluded. The full inclusion list, matched as case-insensitive substrings, was *pharmaceutical*, *prescription*, *medicin*, *medicament*, *therapeutic*, *treatment of*, *treating*, *preparation*, *drug*, *tablet*, *capsule*, *injection*, *ointment*, *antiseptic*, *analgesic*, *antibiotic*, *vaccine*, *remedy*, and *tonic*. A description containing *veterinary* was excluded unless it also contained *human* or *pharmaceutical preparation*.

The Class 005 records were parsed from the 90 files of the Trademark Full-Text XML Data (Applications) annual back file, 2025 edition (files dated December 31, 2025, downloaded February 2026). The back file holds the case files current at its production date, so it includes applications filed in early 2026. Filing dates in the extract run through February 9, 2026, and 1,114 of the 177,337 scored marks were filed in 2026.

As a validation check, we confirmed that well-known names appeared with correct filing years and plausible surprisal values (e.g., PROSCAR filed 1988, WEGOVY 2020). Filing dates preceded FDA approval by several years, as expected.

#### Generic Name Cleaning (International Nonproprietary Names and United States Adopted Names, INN/USAN)

We extracted nonproprietary ingredient names from the same Orange Book new drug application (NDA) data. A combination product lists its ingredients separated by semicolons. We split each such list, and each ingredient entered the generic dataset at its first appearance, alone or in a combination. The list itself was not scored as a name. Splitting added 126 names that the NDA file lists only in combination products (e.g., *sacubitril*, *avibactam*, *ledipasvir*, *elexacaftor*, and *nirmatrelvir*). It also moved 6 names (e.g., caffeine and colchicine) into the augmenting list, because they first appeared in a combination product approved before 1982. Some added names are excipients, vitamins, or sunscreen agents (e.g., glycerin, alcohol, fish oil, vitamin A, and avobenzone), which we kept under the same rules as every other ingredient name. We also excluded radiopharmaceuticals as a category. They were identified by a letter–number isotope designator written with a hyphen or with a space, such as TC-99M, I-123, F-18, and LU 177 (as in LUTETIUM LU 177 DOTATATE). This rule also excluded four non-radiopharmaceutical alphanumeric names (NONOXYNOL-9 and three OMEGA-3 preparations) and two diagnostic agents labeled with stable isotopes (UREA C-13 and hyperpolarized XENON XE-129). Among the ingredients of combination products, it excluded labeled cyanocobalamin (CYANOCOBALAMIN CO-57 and CO-58) and UREA C-13. Finally, we excluded simple element-prefix compounds (e.g., SODIUM CHLORIDE), which are inorganic salts rather than coined names.

We stripped salt and ester suffixes (HYDROCHLORIDE, SODIUM, SULFATE, MESYLATE, and 90 additional terms) right-to-left to recover the base molecule name, so that “DEXAMETHASONE SODIUM PHOSPHATE” became “dexamethasone.” A salt word with a stereochemical prefix, such as S-MALATE or D-TARTRATE, was matched after the prefix was removed, and the two-word counterions DIMETHYL SULFOXIDE and LAURYL SULFATE were stripped as units. We kept acid names as two-word units and normalized them by concatenation, so that “TRANEXAMIC ACID” became “tranexamicacid,” with trigrams computed within each constituent word, as for all multi-word names. We did not strip prodrug ester suffixes (e.g., dipivoxil, cilexetil), because they are part of the INN name and carry their own assigned stems.

#### Same-Day Approvals in the Dynamic Baseline

Names sharing the same approval (or filing) date were not prior to each other. Each name was scored against the names that first appeared on an earlier date, and all names of a date were added to the baseline after all of them had been scored. The nearest-neighbor and neighborhood-count analyses used the same strict date order.

#### Augmented Dynamic Baseline

In the primary analysis, the dynamic baseline of each dataset began with names that were already on the market or on file when the dataset begins. These augmenting names were added to the baseline before the first name was scored. They served only as prior names and were never scored themselves. Names compete with every name on the market, and without them the first names in each dataset would have been compared with very few prior names (28 or fewer for the NME names approved in 1985).

For the Orange Book dataset, the augmenting names were the trade names of Orange Book NDA products approved before 1982 (2,677 product rows, 1,360 unique trade names, 1,198 names after the normalization used for all proprietary names, and 1,030 names after the entries described above were removed). The Orange Book gives no approval date for these products (the date field reads “Approved Prior to Jan 1, 1982”), so they could not be placed in time. All augmenting names of a dataset entered the baseline together, before the first scored name. For the NME dataset, which begins in 1985, the augmenting names were the same pre-1982 names plus the 215 Orange Book NDA trade names approved from 1982 through 1984, 157 of which were not already among the pre-1982 names (1,187 augmenting names). For the generic dataset, the augmenting names were the ingredient names of the pre-1982 NDA products, with combination products split into their ingredients, after the generic-name filters and cleaning described above (733 unique ingredient strings, 650 names). For the USPTO dataset, the augmenting names were Class 005 marks filed before 1980 that passed the same keyword and length filters (9,893 unique names).

The baseline was built in order of first appearance. Every augmenting name was kept, including discontinued products, because their names still occupy name space. A name enters the baseline once, at its first appearance. A later record spelled like a name that had already appeared is not a new name and was not scored. This rule set aside 1 NME name (*Imagent*), 147 Orange Book names (later products that reused a pre-1982 trade name, e.g., *Haldol* and *Lioresal*), 284 generic names (e.g., nitroglycerin and baclofen), and 1,621 USPTO marks. For the NME dataset, every Orange Book NDA trade name first dated 1985 or later (1,618 names) also entered the baseline at its first Orange Book date, so that NME names were compared with all marketed NDA trade names and not only with earlier NME names. The final baselines held 4,126 names (NME), 3,799 (Orange Book), 1,923 (generic), and 187,230 (USPTO). The English baseline does not depend on prior names, so the English-baseline results are the same with and without them.

Under an alternative definition, in which the NME baseline grew only through NME names and augmenting names spelled like a scored name were removed from the augmenting lists instead, the year coefficients were 0.039 (NME), 0.030 (Orange Book), 0.016 (USPTO), and 0.013 (generic).

The augmenting lists have limits. The pre-1982 Orange Book list includes discontinued products that are still listed, but it omits products withdrawn before the Orange Book began in 1980. The NME augmenting names are Orange Book trade names of all NDA products, not only new molecular entities. Biologics licensed before 1985, biosimilars, and other biologics outside the NME compilation are in neither source. The pre-1980 USPTO records consist almost entirely of marks that were registered, because applications abandoned before 1980 are essentially absent from the USPTO file. The augmenting lists are therefore a lower bound on the names in use when each dataset begins.

#### Sensitivity Analysis With an Unaugmented Baseline

We repeated every analysis with an unaugmented dynamic baseline, which contained only names from the same dataset approved earlier and so started empty (Supplementary Table S4, Supplementary Figure S6). The unaugmented analysis scored every name in each dataset, including names already used before the dataset begins (1,322 NME, 2,916 Orange Book, 178,958 USPTO, and 1,557 generic names). Names approved on the first date of a dataset have no prior names, so the regression models omit them (one name in each dataset, two in the USPTO dataset). With the unaugmented baseline, NME top-1 similarity rose from 0.40 in 1987 to 0.56 in 2006, an artifact of the nearly empty early lexicon.

#### Growth of the Baseline (Permutation Null and Fixed References)

Under add-α smoothing (Equation 1 in the main text), a trigram not yet seen costs

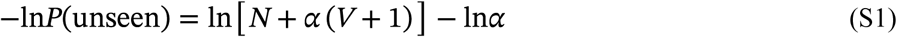

nats, and *N* and *V* grow with the baseline. A growing baseline could therefore raise surprisal over time even if naming did not change. It could also lower it, because a larger baseline leaves fewer of a name’s trigrams unseen. We measured the net effect in three ways (Supplementary Table S5).

First, a permutation null. The scored names of each dataset were randomly reassigned to the observed date slots. The augmenting names, the dates (and so the number of names per year), and, for the NME dataset, the Orange Book trade names at their own dates stayed fixed, and names sharing a date were not prior to each other. Every name was rescored against the augmenting names plus the names placed before it, and the year model (surprisal regressed on year and name length by ordinary least squares) was refit. We ran 5,000 replicates for the NME dataset, 2,000 each for the Orange Book and generic datasets, and 1,000 for the USPTO dataset (all 177,337 marks in each replicate). In every replicate of every dataset the null year coefficient was negative. A growing baseline by itself therefore lowers mean surprisal over time, because the share of unseen trigrams falls faster than the cost of an unseen trigram rises. The observed trends are conservative.

Second, fixed references. Every name was scored against one reference that does not grow, either the final lexicon (augmenting names plus all later names) with the name’s own trigrams removed, or the augmenting names alone. The NME year coefficient was similar with the final lexicon (b = 0.036) and larger with the augmenting names alone (b = 0.054).

Third, a decomposition. Each name’s surprisal is the sum of an unseen-trigram part and a seen-trigram part. The cost of an unseen NME trigram rose only from 11.96 nats (1980s) to 12.97 nats (2020s), while the NME novel fraction rose from 0.072 to 0.147. Holding the cost of an unseen trigram at the value faced by the first scored name left 89% of the NME year coefficient (b = 0.034), 88% for the Orange Book, 91% for generic names, and 98% for the USPTO.

The null and the fixed references show that the trend is not produced by the scoring method. Inventory size and calendar time are nearly collinear, so the same checks apply to the log-inventory model, and they cannot by themselves show that namers responded to crowding.

#### English Baseline Scale and Drug Names in the Word List

The *wordfreq* frequencies are proportions of all English word tokens (they sum to about 1). Entered directly as trigram counts, they give a total count *N* of 6.1, far below the smoothing mass α(*V* + 1) = 1,000 for 10,002 trigram types. Under that scale every trigram has nearly the same probability (an unseen trigram is only 2.8 times less probable than the most common English trigram), so English-baseline surprisal falls in a narrow band (NME 8.95 to 9.22 nats) near its maximum of 9.22 nats, and the year coefficient is small (b = 0.001, R² = 0.067). We therefore entered each word’s frequency as a count per million words, rounded and at least 1 (*N* = 6.2 million). With this scale the English-baseline year coefficient was b = 0.049 (95% CI 0.044 to 0.054, R² = 0.243) for NME names, 0.039 (R² = 0.218) for Orange Book names, 0.019 (R² = 0.169) for USPTO marks, and 0.006 (R² = 0.012, p < .001) for generic names. The English novel fraction does not depend on the count scale, because it depends only on which trigrams occur in the word list.

Of the 50,000 words, 78 (0.05% of the frequency mass) are spelled like an FDA drug name in our data (e.g., *aspirin*, *caffeine*, *Xanax*, *Viagra*, *Valium*). Ten NME names are English-list words (e.g., *Prozac*, *Ambien*, *Allegra*). With these 78 words removed, the NME English novel fraction was unchanged (0.86% in the 1980s and 8.92% in the 2020s, a tenfold rise). The Orange Book 1980s value rose slightly (1.15% to 1.41%), so the Orange Book rise was 5.7-fold instead of 7.0-fold. The English-baseline year coefficient for NME names was also unchanged (b = 0.049).

#### Brand and Generic Neighborhood Counts at Equal Lexicon Size

The brand (Orange Book) lexicon is larger than the generic lexicon (1,030 augmenting names and 2,769 names against 650 augmenting names and 1,273 names), and a larger lexicon offers more potential neighbors. We drew random subsets of the brand augmenting names and scored names of the generic sizes, kept their chronological order, recomputed each brand name’s count of prior names within Levenshtein distance 3 (names written with single spaces between words), and refit the length-adjusted brand–generic gap against the unchanged generic counts (500 replicates). The mean brand count fell from 2.99 to 1.60 (generic 0.60), and the gap fell from 1.38 to 0.51 fewer neighbors for generic names (95% of replicates between 0.40 and 0.63, all replicates negative), so nearly two-thirds of the gap (63%) reflects lexicon size. Per 1,000 prior names, brand names had 1.47 neighbors within distance 3 and generic names 0.51. Name length was associated with neighborhood count (each additional character predicted 0.48 fewer neighbors for brand names and 0.07 fewer for generic names, both p < .001), and adjusting for length reduced the gap from 2.39 to 1.38.

#### Sensitivity to the Smoothing Constant

We rescored the NME names against the augmented dynamic drug name baseline with α = 0.01 and α = 1.0, in addition to the α = 0.1 used in the main analysis, and refit the year model of Table 3 each time (Supplementary Table S3). The year coefficient was positive and significant (p < .001) at all three values. The coefficient became smaller as α increased (b = 0.042 to 0.032), and the variance explained became larger (R² = 0.274 to 0.420).

### Supplementary Tables

**Supplementary Table S1.**
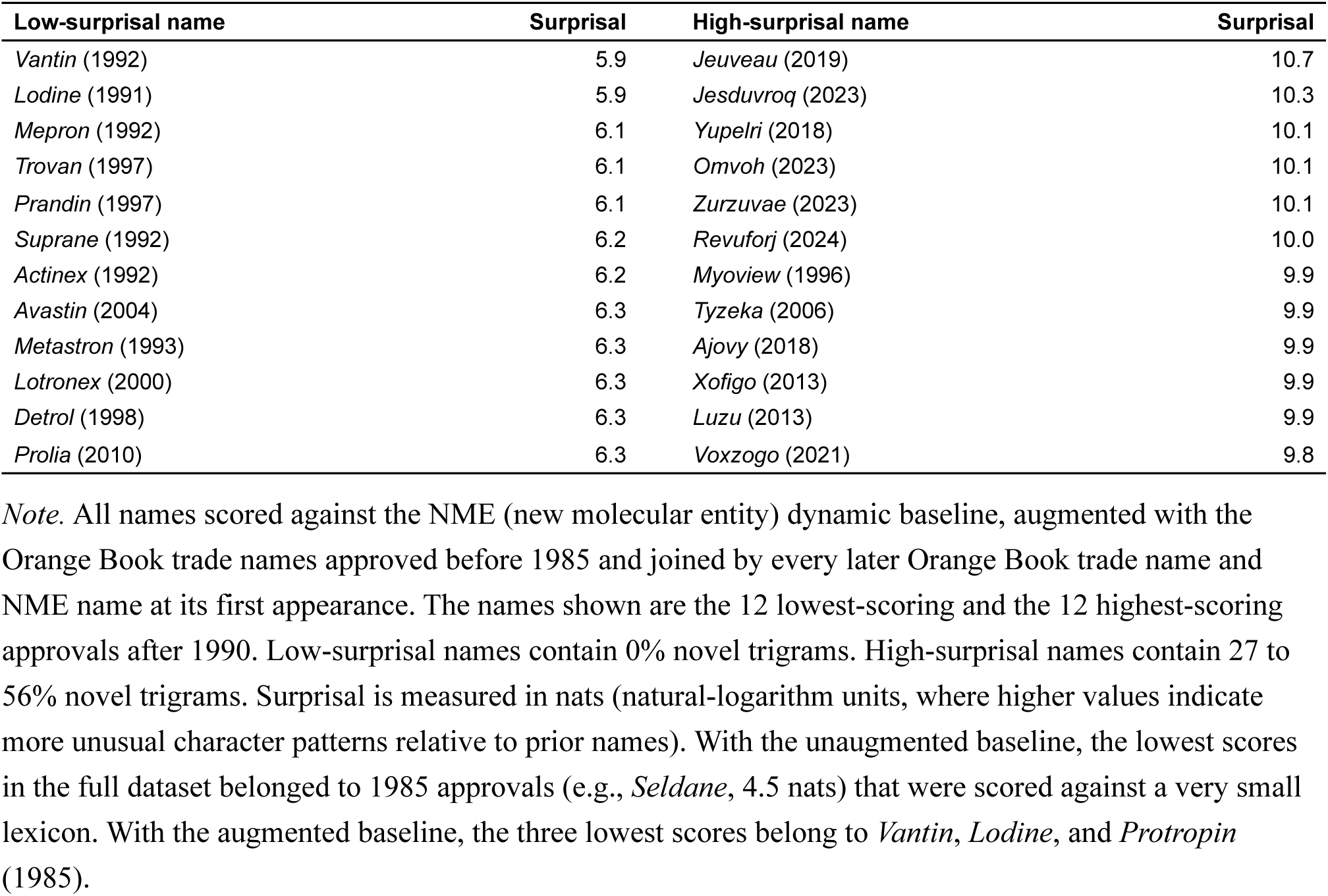
NME drug names at the extremes of the trigram surprisal distribution (approvals after 1990).

**Supplementary Table S2.**
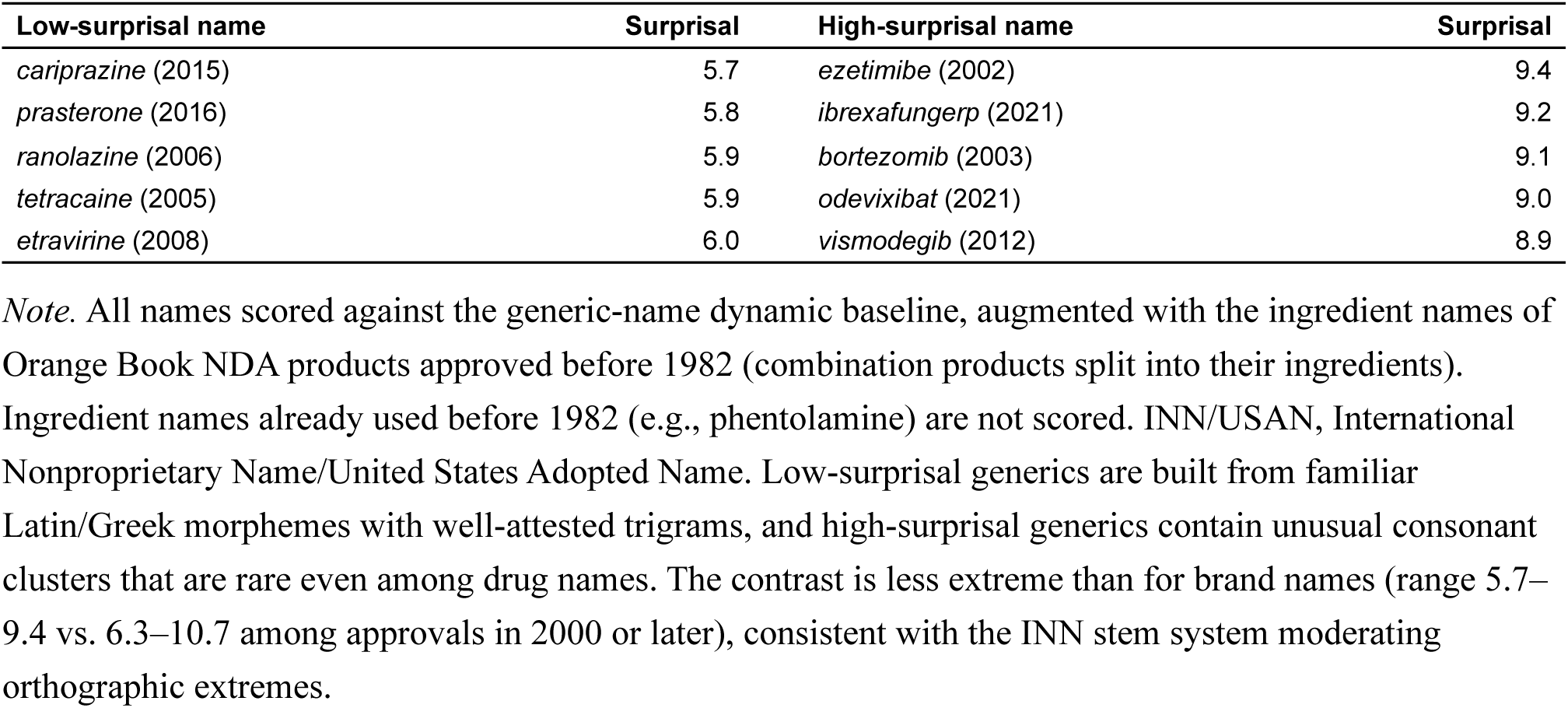
Generic (INN/USAN) drug names at the extremes of the trigram surprisal distribution (approvals in 2000 or later).

**Supplementary Table S3.** Sensitivity of the NME year coefficient to the add-α smoothing constant.

| Smoothing constant ($\alpha$) | b (nats/year) | 95% CI | SE | p | R<sup>2</sup> |
| --- | --- | --- | --- | --- | --- |
| 0.01 | 0.042 | [0.039, 0.046] | 0.002 | < .001 | 0.274 |
| 0.1 (main analysis) | 0.038 | [0.035, 0.041] | 0.001 | < .001 | 0.338 |
| 1.0 | 0.032 | [0.030, 0.034] | 0.001 | < .001 | 0.420 |

*Note.* Coefficient for approval year in the NME year model (drug baseline trigram surprisal regressed on approval year and name length, ordinary least squares with HC3 robust standard errors, $N = 1,321$ ), refit after every name was rescored with each smoothing constant against the augmented baseline. The $\alpha = 0.1$ row repeats Table 3. NME, new molecular entity. b, regression coefficient. CI, confidence interval. SE, standard error.

**Supplementary Table S4.** Main results with the augmented (primary) and the unaugmented dynamic baseline.

*Panel A. Trigram surprisal (Tables 1 and 2).*
| Dataset | Augmenting names | Mean surprisal, 1980s<br>(augmented / unaugmented) | Mean surprisal, 2020s<br>(augmented / unaugmented) | Increase (augmented / unaugmented) | Year b [95% CI], augmented | Year b [95% CI], unaugmented | R<sup>2</sup> (augmented / unaugmented) |
| --- | --- | --- | --- | --- | --- | --- | --- |
| NME | 1,187 | 7.21 / 6.97 | 8.50 / 8.24 | 18% / 18% | 0.038 [0.035, 0.041] | 0.035 [0.032, 0.038] | 0.338 / 0.320 |
| Orange Book | 1,030 | 7.19 / 7.17 | 8.34 / 8.25 | 16% / 15% | 0.031 [0.029, 0.033] | 0.028 [0.027, 0.030] | 0.285 / 0.275 |
| USPTO | 9,893 | 7.24 / 7.27 | 7.75 / 7.75 | 7% / 7% | 0.016 [0.016, 0.017] | 0.016 [0.016, 0.017] | 0.148 / 0.149 |
| Generic (INN/USAN) | 650 | 6.88 / 6.77 | 7.29 / 7.20 | 6% / 6% | 0.011 [0.008, 0.013] | 0.011 [0.009, 0.013] | 0.063 / 0.072 |

*Panel B. Nearest-neighbor similarity, novel fraction, and neighborhood counts.*
| Measure | Augmented | Unaugmented |
| --- | --- | --- |
| NME prior names behind a 1985 score / the last 2024 score | 1,194–1,254 / 4,034 | 0–28 / 1,320 |
| NME closest-name similarity, 1987 (5-year rolling mean) | 0.61 | 0.40 |
| NME closest-name similarity, peak (year) | 0.62 (1992) | 0.56 (2006) |
| NME closest-name similarity, 2005 / 2024 | 0.59 / 0.54 | 0.55 / 0.52 |
| NME 10-closest similarity, peak (year) / 2024 | 0.52 (1992) / 0.47 | 0.45 (2006) / 0.44 |
| NME drug baseline novel fraction, 1985–1989 / 2020s | 0.07 / 0.15 | 0.55 / 0.23 |
| Orange Book closest-name similarity, peak (year) / 2020s | 0.62 (1988) / 0.56 | 0.59 (2006) / 0.55 |
| Orange Book prior names within distance 3, 1980s / 2000s / 2020s | 2.84 / 3.71 / 2.13 | 0.70 / 2.71 / 1.77 |
| Orange Book year coefficient for neighbor count (p) | –0.045 (< .001) | 0.015 (< .001) |
| Orange Book squared-year coefficient (p), R<sup>2</sup>, fitted peak | –0.0034 (< .001), 0.19, 1998 | –0.0039 (< .001), 0.18, 2006 |
| Generic prior names within distance 3, 1980s / 2010s / 2020s | 0.64 / 0.60 / 0.58 | 0.27 / 0.59 / 0.55 |
| Generic year coefficient for neighbor count (p) | 0.000 (.94) | 0.009 (< .001) |
| Generic minus brand neighbor count, length-adjusted | -1.38 | -0.78 |

*Panel C. Year coefficient with standard errors clustered by approval (filing) year.*
| Dataset | b, augmented | SE, HC3 / clustered, augmented | 95% CI, clustered, augmented | b, unaugmented | SE, HC3 / clustered, unaugmented | 95% CI, clustered, unaugmented |
| --- | --- | --- | --- | --- | --- | --- |
| NME | 0.038 | 0.0015 / 0.0021 | [0.034, 0.042] | 0.035 | 0.0014 / 0.0022 | [0.031, 0.039] |
| Orange Book | 0.031 | 0.0010 / 0.0018 | [0.027, 0.034] | 0.028 | 0.0009 / 0.0018 | [0.025, 0.032] |
| USPTO | 0.016 | 0.0002 / 0.0010 | [0.015, 0.018] | 0.016 | 0.0002 / 0.0010 | [0.014, 0.018] |
| Generic (INN/USAN) | 0.011 | 0.0013 / 0.0014 | [0.008, 0.013] | 0.011 | 0.0011 / 0.0010 | [0.009, 0.013] |

**Supplementary Table S5.** Growth of the baseline (permutation null, fixed references, and decomposition of the year coefficient).

*Panel A. Permutation null (names randomly reassigned to the observed dates).*
| Dataset | Replicates | Observed b | Null b, mean [2.5th, 97.5th percentile] | Largest null b | Observed rise, 1980s to 2020s (nats) | Null rise, mean (nats) |
| --- | --- | --- | --- | --- | --- | --- |
| NME | 5,000 | 0.038 | -0.010 [-0.014, -0.007] | -0.004 | 1.29 | -0.38 |
| Orange Book | 2,000 | 0.031 | -0.007 [-0.009, -0.005] | -0.002 | 1.15 | -0.28 |
| USPTO | 1,000 | 0.0164 | -0.0020 [-0.0024, -0.0017] | -0.0015 | 0.51 | -0.13 |
| Generic (INN/USAN) | 2,000 | 0.011 | -0.009 [-0.012, -0.007] | -0.005 | 0.41 | -0.37 |

*Panel B. Year coefficient against references that do not grow.*
| Dataset | Dynamic (primary), b [95% CI], R<sup>2</sup> | Final lexicon, leave-one-out, b [95% CI], R<sup>2</sup> | Augmenting names only, b [95% CI], R<sup>2</sup> |
| --- | --- | --- | --- |
| NME | 0.038 [0.035, 0.041], 0.338 | 0.036 [0.033, 0.038], 0.378 | 0.054 [0.051, 0.057], 0.454 |
| Orange Book | 0.031 [0.029, 0.033], 0.285 | 0.029 [0.027, 0.030], 0.313 | 0.043 [0.041, 0.045], 0.370 |
| USPTO | 0.0164 [0.0161, 0.0167], 0.148 | 0.0146 [0.0143, 0.0149], 0.157 | 0.0236 [0.0233, 0.0240], 0.137 |
| Generic (INN/USAN) | 0.011 [0.008, 0.013], 0.063 | 0.012 [0.010, 0.014], 0.080 | 0.033 [0.030, 0.037], 0.237 |

*Panel C. Year coefficient of each part of surprisal (observed).*
| Dataset | Unseen-trigram part, b | Seen-trigram part, b | Surprisal with the unseen-trigram cost held fixed, b (share of total b) |
| --- | --- | --- | --- |
| NME | 0.027 | 0.011 | 0.034 (89%) |
| Orange Book | 0.017 | 0.014 | 0.027 (88%) |
| USPTO | -0.0014 | 0.0178 | 0.0160 (98%) |
| Generic (INN/USAN) | -0.009 | 0.020 | 0.010 (91%) |

*Note.* Year models regress drug baseline trigram surprisal on approval (filing) year and name length. In Panel A, the scored names were shuffled over the observed date slots, the augmenting names and (for the NME dataset) the Orange Book trade names stayed at their own dates, every name was rescored, and the model was refit by ordinary least squares. Every null coefficient in every dataset was negative, and no replicate reached the observed coefficient (empirical one-sided $p = 1/(\text{replicates} + 1)$ ). Rise is the 2020s mean minus the 1980s mean. In Panel B, the final lexicon is all augmenting names and all later names, with the scored name's own trigrams removed. Panels B and C use HC3 robust standard errors. In Panel C, the two parts add up to the total coefficient, and the last column holds the cost of an unseen trigram at the value faced by the first scored name. USPTO values are given to four decimals because its coefficients are small. NME, new molecular entity. USPTO, United States Patent and Trademark Office. INN/USAN, International Nonproprietary Name/United States Adopted Name. b, regression coefficient. CI, confidence interval.

**Supplementary Table S6.** Year coefficient for single-word names.

| **Dataset** | **All names, b [95% CI], R<sup>2</sup> (N)** | **Single-word names, full baseline, b [95% CI], R<sup>2</sup> (N)** | **Single-word names, single-word baseline, b [95% CI], R<sup>2</sup>** |
| --- | --- | --- | --- |
| NME | 0.038 [0.035, 0.041], 0.338<br>(1,321) | 0.039 [0.036, 0.042], 0.345<br>(1,291) | 0.038 [0.035, 0.041], 0.344 |
| Orange Book | 0.031 [0.029, 0.033], 0.285<br>(2,769) | 0.033 [0.031, 0.035], 0.301<br>(2,224) | 0.032 [0.030, 0.035], 0.295 |
| USPTO | 0.0164 [0.0161, 0.0167], 0.148<br>(177,337) | 0.0230 [0.0226, 0.0234], 0.122<br>(115,594) | 0.0219 [0.0215, 0.0223], 0.112 |
| Generic (INN/USAN) | 0.011 [0.008, 0.013], 0.063<br>(1,273) | 0.011 [0.008, 0.014], 0.060<br>(1,164) | 0.010 [0.007, 0.012], 0.054 |

*Note.* Year models regress drug baseline trigram surprisal on approval (filing) year and name length (ordinary least squares, HC3 robust standard errors). “Full baseline” scores single-word names against the primary augmented baseline, which includes multi-word names. “Single-word baseline” rescores them against augmenting names and earlier names that are single words. All year coefficients have $p < .001$ . NME, new molecular entity. USPTO, United States Patent and Trademark Office. INN/USAN, International Nonproprietary Name/United States Adopted Name. b, regression coefficient. CI, confidence interval.

**Supplementary Table S7.** Post-2000 NME approvals whose closest prior FDA-approved name had a normalized Levenshtein similarity of 0.70 or more.

| **New name**<br>**(approval year)** | **Closest prior**<br>**name (first year,**<br>**source)** | **Similarity** | **Edits** | **2026 status, new**<br>**name** | **2026 status,**<br>**prior name** |
| --- | --- | --- | --- | --- | --- |
| *Ventavis* (2004) | *Vertavis* (before 1982, Orange Book) | 0.88 | 1 | Discontinued | Discontinued |
| *Besivance* (2009) | *Kepivance* (2004, NME BLA) | 0.78 | 2 | Marketed | Marketed |
| *Vitrase* (2004) | *Vitrasert* (1996, Orange Book) | 0.78 | 2 | Marketed | Discontinued |
| *Ella* (2010) | *Emla* (1992, Orange Book) | 0.75 | 1 | Marketed | Discontinued |
| *Eovist* (2008) | *Renovist* (before 1982, Orange Book) | 0.75 | 2 | Marketed | Discontinued |
| *Faslodex* (2002) | *Casodex* (1995, NME NDA) | 0.75 | 2 | Marketed | Marketed |
| *Dalvance* (2014) | *Dalmane* (before 1982, Orange Book) | 0.75 | 2 | Marketed | Discontinued |
| *Bridion* (2015) | *Tridione* (before 1982, Orange Book) | 0.75 | 2 | Marketed | Discontinued |
| *Septocaine* (2000) | *Sensorcaine* (before 1982, Orange Book) | 0.73 | 3 | Marketed | Marketed |
| *Folotyn* (2009) | *Solodyn* (2006, Orange Book) | 0.71 | 2 | Marketed | Discontinued |
| *Rapivab* (2014) | *Raptiva* (2003, NME BLA) | 0.71 | 2 | Marketed | Discontinued |
| *Harvoni* (2014) | *Darvon* (before 1982, Orange Book) | 0.71 | 2 | Marketed | Discontinued |
| *Solosec* (2017) | *Losec* (1989, NME NDA) | 0.71 | 2 | Marketed | Not listed |
| *Stendra* (2012) | *Stelara* (2009, NME BLA) | 0.71 | 2 | Marketed | Marketed |
| *Edurant* (2011) | *Duract* (1997, NME NDA) | 0.71 | 2 | Marketed | Not listed |
| *Vyondys 53* (2019) | *Exondys 51* (2016, NME NDA) | 0.71 | 2 | Marketed | Marketed |
| *Halaven* (2010) | *Talacen* (1982, Orange Book) | 0.71 | 2 | Marketed | Discontinued |
| *Gemtesa* (2020) | *Genesa* (1997, NME NDA) | 0.71 | 2 | Marketed | Discontinued |
| *Xuriden* (2015) | *Doriden* (before 1982, Orange Book) | 0.71 | 2 | Marketed | Discontinued |
| *Coartem* (2009) | *Cortef* (before 1982, Orange Book) | 0.71 | 2 | Marketed | Marketed |

| New name<br>(approval year) | Closest prior<br>name (first year,<br>source) | Similarity | Edits | 2026 status, new<br>name | 2026 status,<br>prior name |
| --- | --- | --- | --- | --- | --- |
| *Sabril* (2009) | *Sandril* (before 1982, Orange Book) | 0.71 | 2 | Marketed | Discontinued |
| *Foradil* (2001) | *Toradol* (1989, NME NDA) | 0.71 | 2 | Discontinued | Discontinued |
| *Pristiq* (2008) | *Priftin* (1998, NME NDA) | 0.71 | 2 | Marketed | Marketed |
| *Mircera* (2007) | *Mirena* (2000, Orange Book) | 0.71 | 2 | Marketed | Marketed |
| *Altabax* (2007) | *Atarax* (before 1982, Orange Book) | 0.71 | 2 | Discontinued | Discontinued |
| *Veregen* (2006) | *Verelan* (1990, Orange Book) | 0.71 | 2 | Marketed | Marketed |
| *Chantix* (2006) | *Cantil* (before 1982, Orange Book) | 0.71 | 2 | Marketed | Discontinued |
| *Dacogen* (2006) | *Desogen* (1992, NME NDA) | 0.71 | 2 | Discontinued | Discontinued |
| *Avastin* (2004) | *Fastin* (before 1982, Orange Book) | 0.71 | 2 | Marketed | Discontinued |
| *Benicar* (2002) | *Xenical* (1999, NME NDA) | 0.71 | 2 | Marketed | Marketed |
| *Orfadin* (2002) | *Rifadin* (before 1982, Orange Book) | 0.71 | 2 | Marketed | Marketed |
| *Kineret* (2001) | *Sinemet* (before 1982, Orange Book) | 0.71 | 2 | Marketed | Marketed |
| *Amondys 45* (2021) | *Omontys* (2012, NME NDA) | 0.71 | 2 | Marketed | Discontinued |

*Note.* Similarity is $1 - \text{edit distance} / \text{length of the longer name}$ , with the words of a multi-word name joined by a single space (Equation 3 in the main text). Prior names include the augmenting names and every earlier NME name and Orange Book NDA trade name. NME, new molecular entity compilation (NDA or BLA). Status is taken from the FDA Orange Book Products file (February 2026) for NDA products and the Purple Book (March 2026) for biologics. A name is listed as marketed if any of its products is listed as prescription or over-the-counter, and as discontinued if all of its products are listed as discontinued. The files give no discontinuation dates, so a discontinued prior product might still have been marketed when the new name was approved. Not listed, name not found in the 2026 Orange Book (*Losec* was renamed *Prilosec*, and *Duract* was withdrawn in 1998). Of the 33 pairs, both products are marketed in 12, and the prior product is discontinued in 19 and not listed in 2.

**Supplementary Table S8.** Names containing letter sequences absent from English, by decade.

| **Dataset** | **Decade** | **N names** | **Contains a trigram**<br>**absent from English** | **Contains *q* not**<br>**followed by *u*** |
| --- | --- | --- | --- | --- |
| NME | 1980s | 122 | 6.6% | 0.0% |
| NME | 1990s | 337 | 8.0% | 0.0% |
| NME | 2000s | 250 | 17.2% | 0.8% |
| NME | 2010s | 370 | 24.9% | 4.6% |
| NME | 2020s | 242 | 49.6% | 7.4% |
| Orange Book | 1980s | 396 | 9.8% | 0.8% |
| Orange Book | 1990s | 576 | 10.1% | 0.5% |
| Orange Book | 2000s | 582 | 16.5% | 1.4% |
| Orange Book | 2010s | 737 | 24.6% | 3.8% |
| Orange Book | 2020s | 478 | 47.5% | 7.1% |
| USPTO | 1980s | 6,842 | 7.9% | 0.2% |
| USPTO | 1990s | 18,726 | 8.3% | 0.6% |
| USPTO | 2000s | 40,693 | 11.9% | 1.0% |
| USPTO | 2010s | 54,792 | 15.9% | 1.8% |
| USPTO | 2020s | 56,284 | 22.7% | 2.5% |
| Generic (INN/USAN) | 1980s | 208 | 16.3% | 0.0% |
| Generic (INN/USAN) | 1990s | 312 | 16.3% | 0.0% |
| Generic (INN/USAN) | 2000s | 246 | 15.0% | 0.0% |
| Generic (INN/USAN) | 2010s | 295 | 13.2% | 0.0% |
| Generic (INN/USAN) | 2020s | 212 | 17.9% | 0.0% |

*Note.* A trigram is absent from English if it occurs in none of the 50,000 most common English words of the English baseline (boundary trigrams included, so a name can qualify by how it begins or ends). The NME 1980s cover 1985–1989. The 2020s cover 2020–2024 for NME, 2020–2025 for generic names, and 2020 through early 2026 for the Orange Book and USPTO datasets. The NME names with a *q* not followed by *u* (37, all approved in 2008 or later) are listed in the Zenodo deposit (results\_primary\_v12/weird\_names/q\_without\_u\_nme.csv).

### Supplementary Figure Legends

**Supplementary Figure S1.**
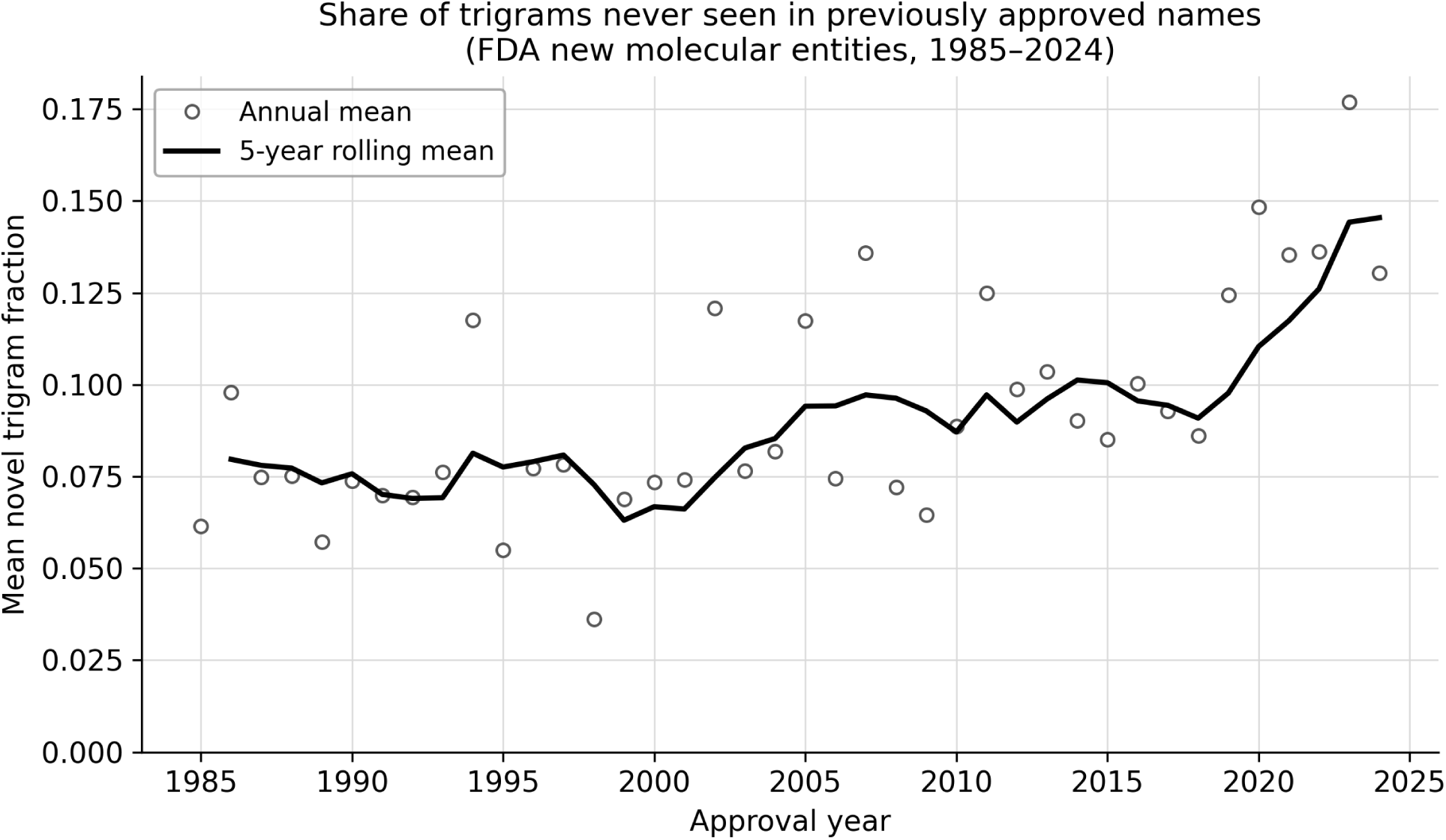
Mean drug baseline novel trigram fraction by approval year (NME dataset). Open circles show annual means, and the line shows the 5-year rolling mean. The novel fraction is the proportion of a name’s character trigrams that had zero count in the dynamic baseline at the time of approval (the Orange Book trade names approved before 1985, plus later Orange Book trade names and NME names from their first appearance).

**Supplementary Figure S2.**
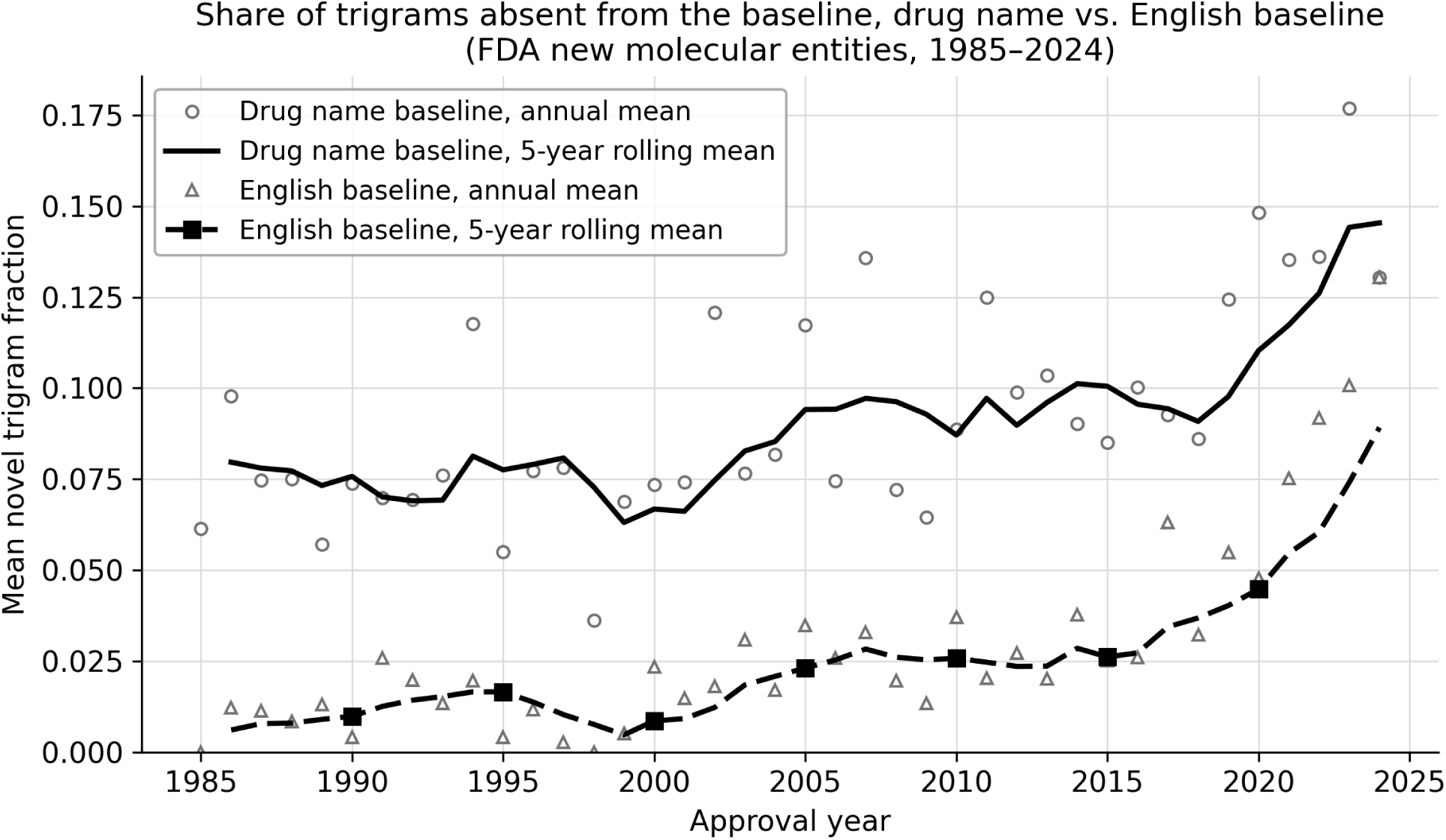
Mean novel trigram fraction by approval year, comparing drug baseline and English-baseline models (NME dataset). Open markers show annual means, and lines show 5-year rolling means. The English novel fraction rises from near zero in the 1980s to approximately 13% by 2024.

**Supplementary Figure S3.**
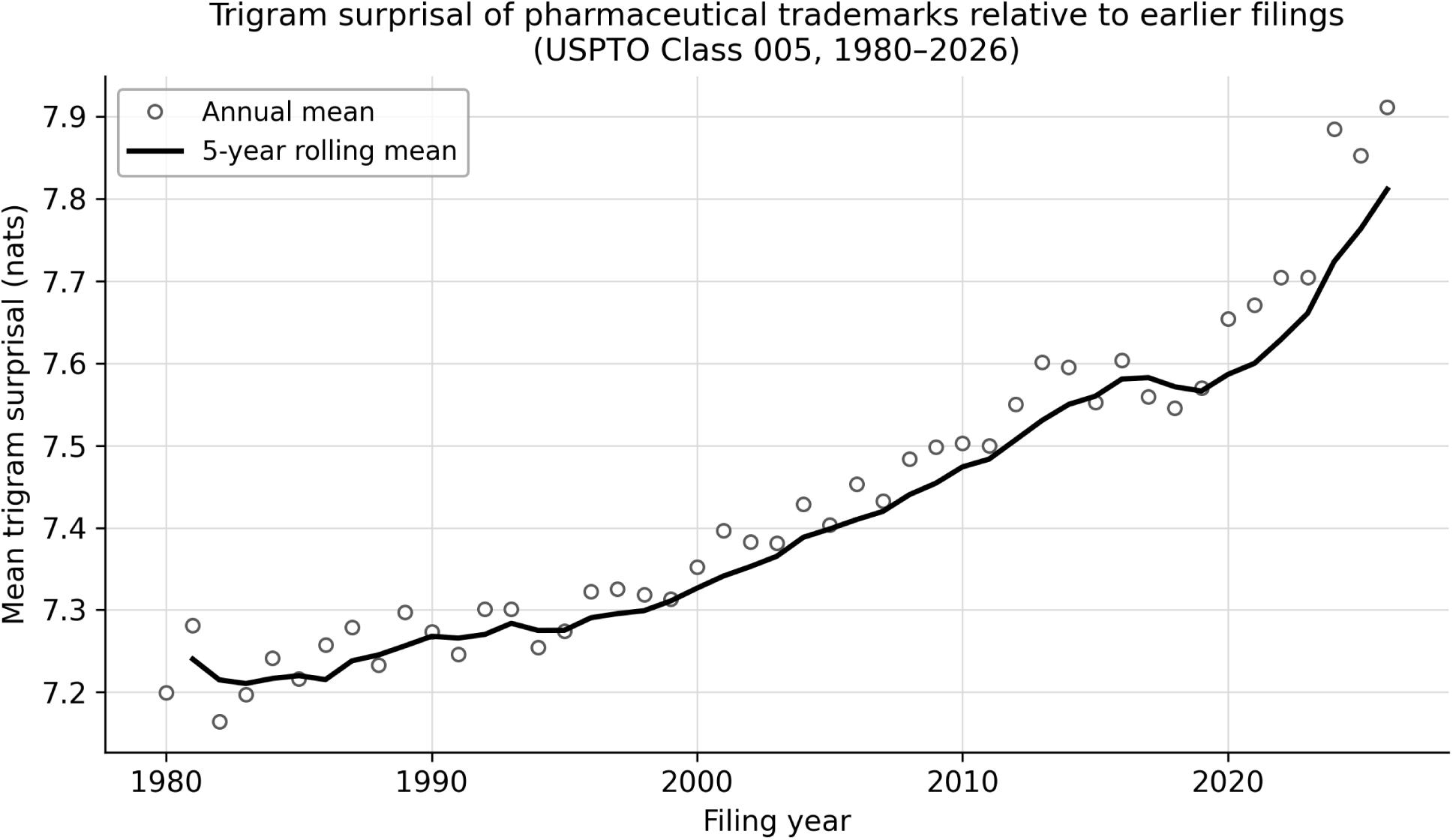
Mean drug baseline character trigram surprisal by filing year (USPTO Class 005 pharmaceutical trademarks, 1980–2026). Each point is the annual mean, and the line shows a 5-year rolling average. Prior names include the Class 005 marks filed before 1980. The steady upward trend replicates the NME and Orange Book findings at 134-fold scale.

**Supplementary Figure S4.**
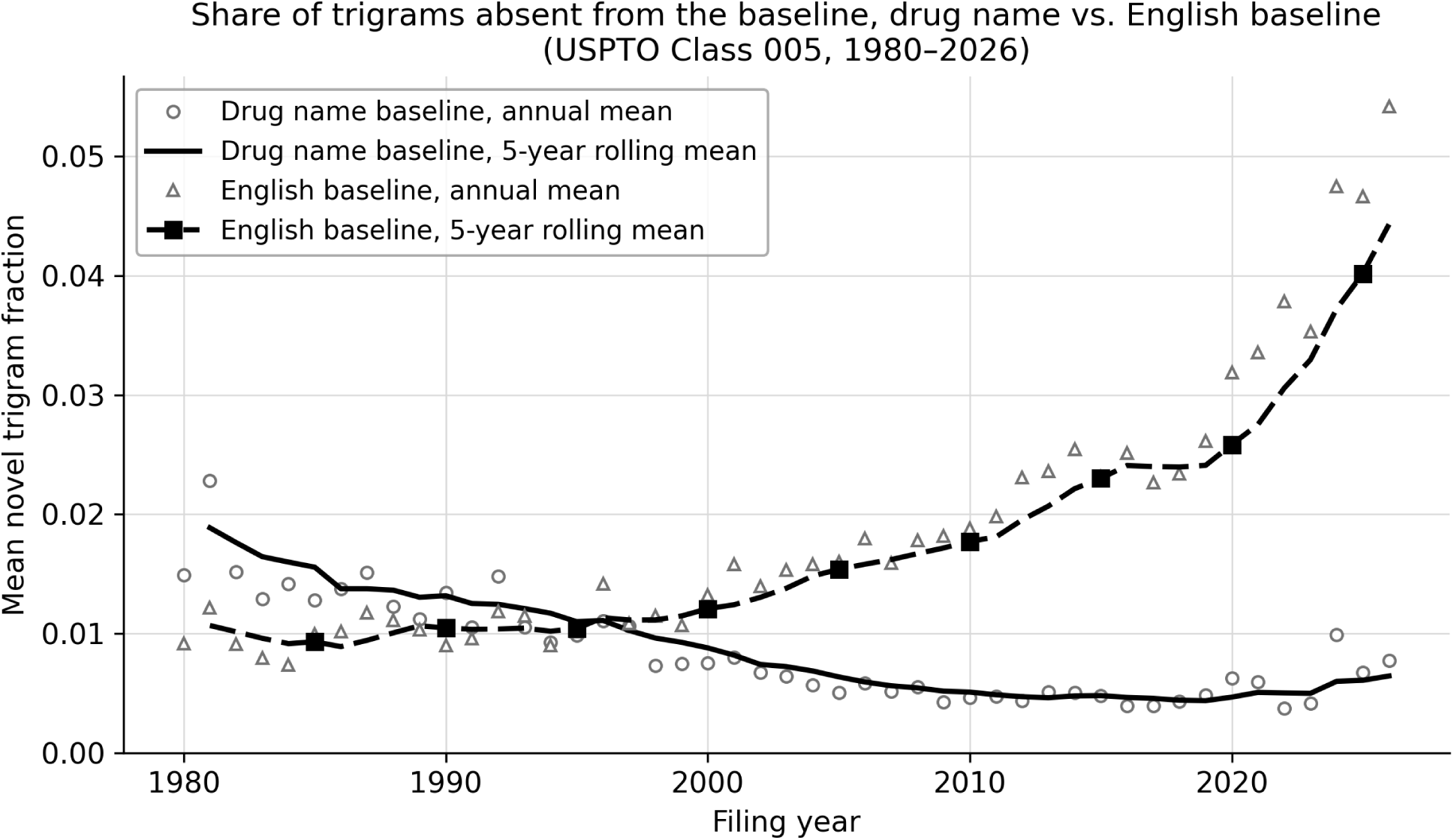
Mean novel trigram fraction by filing year, comparing drug baseline and English-baseline models (USPTO Class 005). Open markers show annual means, and lines show 5-year rolling means. The English novel fraction rises from 1.0% in the 1980s to 3.9% in the 2020s.

**Supplementary Figure S5.**
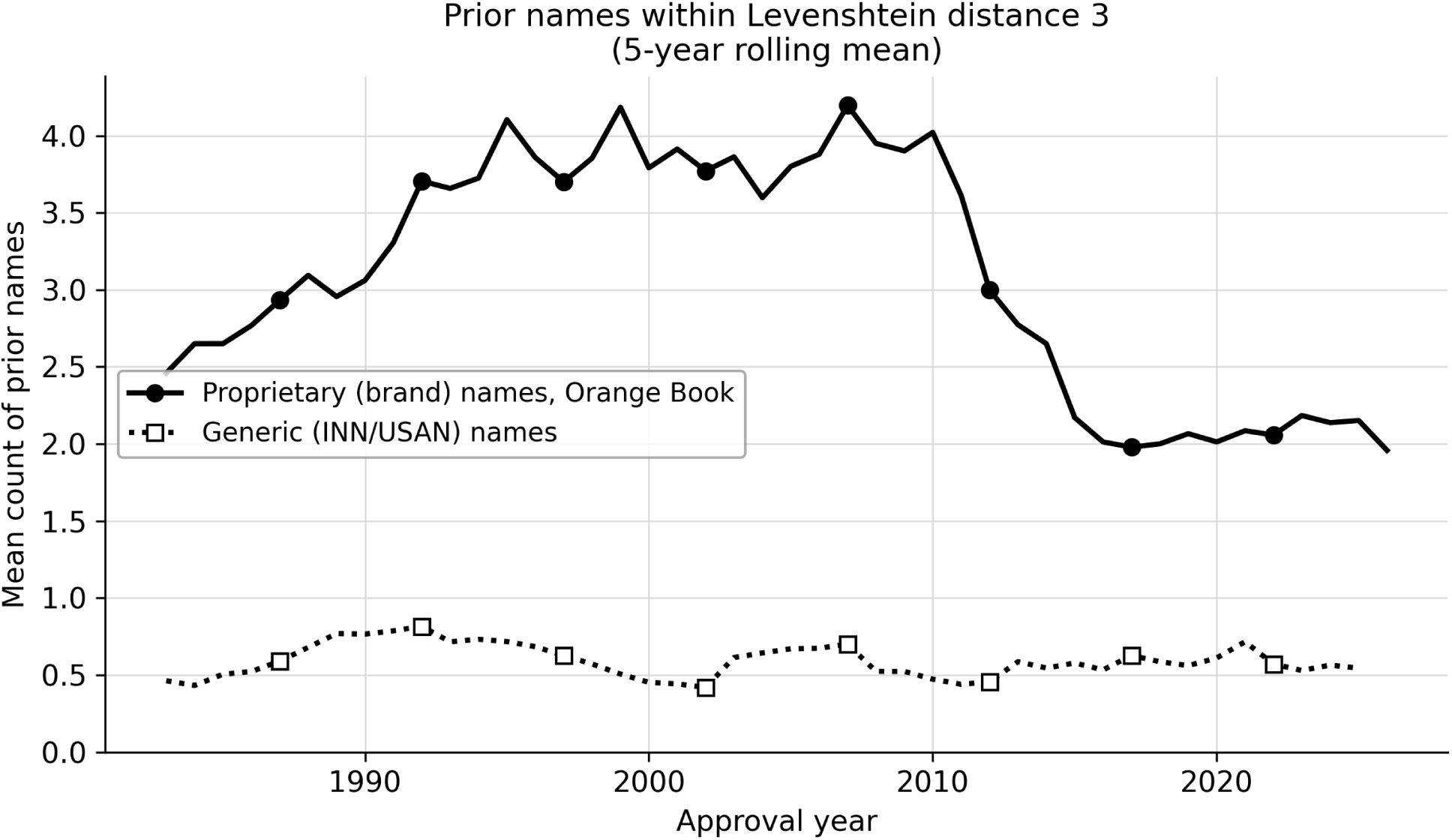
Mean count of prior names within Levenshtein distance 3 by approval year for proprietary (Orange Book brand) and generic (INN/USAN) names, 1982–2026 (5-year rolling averages). Prior names include names approved before 1982. Brand names show an inverted-U pattern, rising to a plateau from the mid-1990s to about 2010 and then falling, while generic names remain relatively flat.

**Supplementary Figure S6.**
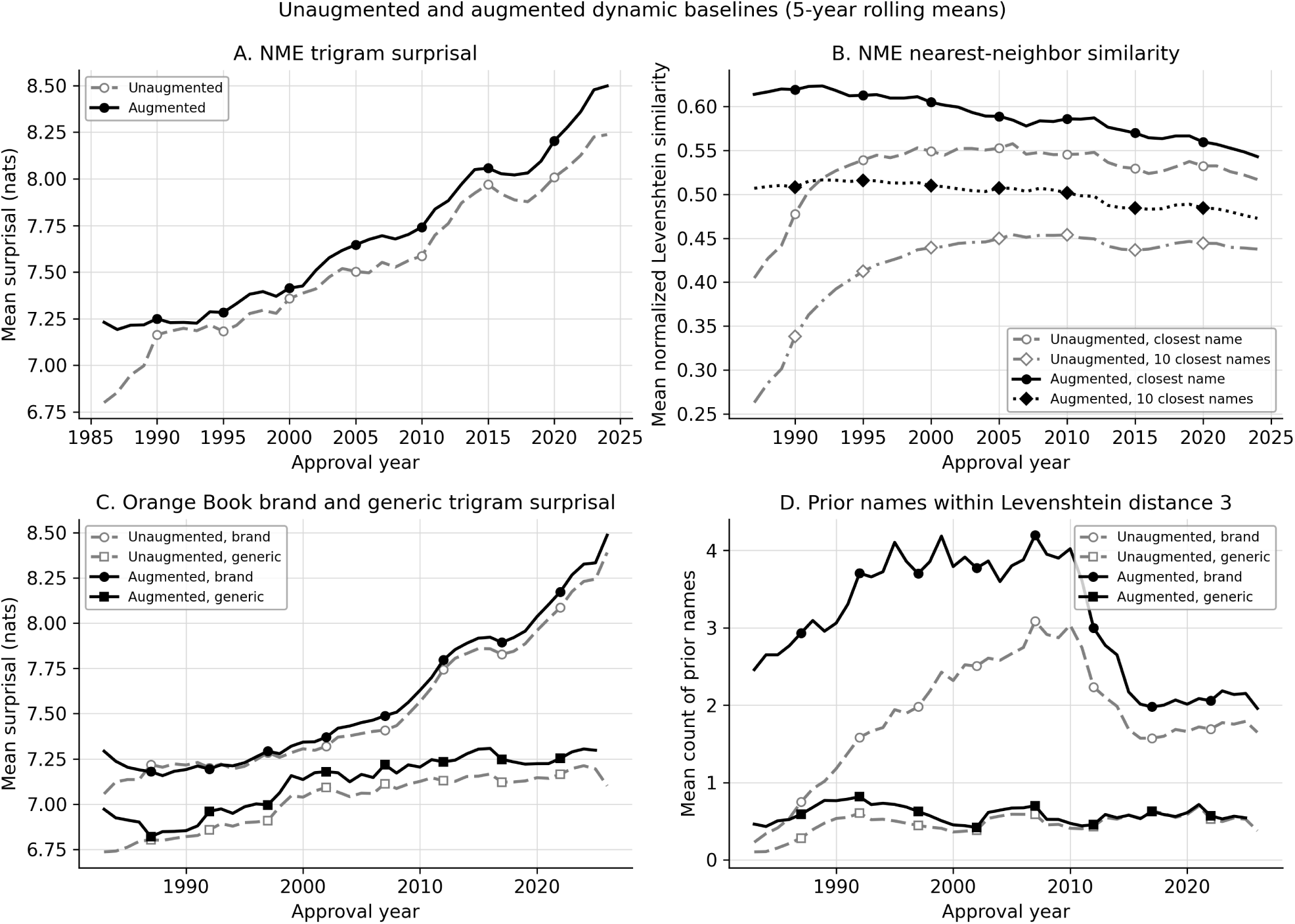
Augmented and unaugmented dynamic baselines compared (5-year rolling means). (A) Mean trigram surprisal of NME names. (B) Mean similarity of each NME name to its closest prior name and to its 10 closest prior names. (C) Mean trigram surprisal of Orange Book brand names and generic names. (D) Mean number of prior names within Levenshtein distance 3 for Orange Book brand names and generic names. Gray dashed lines with open markers (“Unaugmented”) use the unaugmented baseline (prior names from the same dataset only). Black solid lines with filled markers (“Augmented”) use the primary augmented baseline, which also includes names on the market before each dataset begins. With the augmented baseline, the surprisal trends keep their shape, and the early rise in nearest-neighbor similarity disappears.

